# Pan-fibrotic gene expression signature in major chronic diseases by integrative bulk and single-cell transcriptomic analyses

**DOI:** 10.1101/2025.10.07.25337472

**Authors:** Mayra Alejandra Jaimes Campos, Rafael Stroggilos, Joost-Peter Schanstra, Antonia Vlahou, Manon Brunet, Jean-Christophe Jonas, Mohammed Bensellam, Harald Mischak, Agnieszka Latosinska

## Abstract

**Background:** Fibrosis is a common pathological endpoint across multiple chronic diseases affecting organs, including the liver, kidney, and heart. Despite its prevalence and clinical burden, there are currently no robust validated molecular signatures that capture shared fibrotic mechanisms across organ systems. Herein, we present a comprehensive, human-based, cross-organ transcriptomic analysis that identifies a conserved pan-fibrotic gene expression signature.

**Methods:** A comprehensive, cross-platform, and multi-layer transcriptomic analysis of human fibrosis was conducted using microarray, bulk RNA-seq, and single-nucleus RNA-seq data. The study integrated data from 1859 human tissue samples across liver, kidney, and heart fibrosis. The organ-specific and pan-fibrotic gene signatures across organs were initially defined based on a discovery cohort microarray data (n = 1051) and validation cohort (n = 325), while two RNA-seq datasets (n = 414) and four integrated liver snRNA-seq datasets (n = 69) were used as independent validation cohorts. Findings were evaluated using differential expression, pathway enrichment, and protein interaction analyses.

**Findings:** We identified a conserved, pan-fibrotic transcriptional signature comprising 497 genes consistently associated with fibrosis across liver, kidney, and heart tissues. From these, 23 hallmark genes were shortlisted, including both known fibrosis markers (e.g., *CCL5*, *SERPINE2*, *THBS2*, *COL5A1*, *VEGFC*, *SOX4*, among others) and novel candidates (e.g., *SYT11*, *CRIP1*, *PLA2G4C*, *ARHGEF2*, *CA2*, *ELL2, MT1X,* and *MT1E*). Validation in two independent RNA-seq cohorts confirmed 19 genes, with expression levels of 15 significantly correlated with fibrosis severity. Single-nucleus RNA-seq analysis further refined the signature to 11 robustly validated genes exhibiting distinct, cell-type-specific expression profiles. Pathway analysis highlighted significant activation of extracellular matrix remodeling and inactivation of metabolic and ion-homeostasis pathways. The interaction network demonstrated strong interconnectivity among these hallmark genes within key fibrotic modules.

**Interpretation:** The pan-fibrotic gene expression signature offers potential as a cross-organ biomarker set for fibrosis progression and may support the development of broad-spectrum anti-fibrotic therapies.

## Introduction

Fibrosis is a pathological process characterized by disrupted extracellular matrix (ECM) homeostasis, leading to excessive accumulation of ECM components. The progression of fibrosis is a highly dynamic and multi-factorial process. It represents the final common pathway in a wide range of chronic diseases affecting vital organs, including the liver, kidney, lung, and heart. Progressive fibrosis results in tissue remodeling, organ dysfunction, and ultimately, organ failure, representing a substantial societal and economic burden worldwide (*1*).

Despite significant unmet needs, the development of effective anti-fibrotic therapies remains scarce by the lack of early diagnostic and prognostic biomarkers, as well as a limited number of validated molecular targets (*2*). Furthermore, understanding the complex molecular mechanisms underlying fibrosis initiation and progression is essential for developing new treatment strategies (*1*, *3*, *4*). While tissue-specific mechanisms have been extensively studied, the processes that sustain chronic fibrosis, believed to follow conserved molecular principles across organs, remain poorly understood (*2*). Therefore, identifying common molecular signatures and core signaling pathways in humans with cross-tissue relevance, continues to be a research priority.

Advances in next-generation sequencing and transcriptomic profiling have led to the characterization of numerous fibrosis-associated genes (e.g., *TGF-β1*, *COL1A1*, *LOXL2*, *miR-21*, among others), uncovering critical drivers and key mechanisms (*1*, *5–7*). Furthermore, several signaling pathways have been implicated in the development and progression of fibrosis, and some have been described in multiple tissues and disease contexts, including among others, ECM remodelling, TGFβ–SMAD, connective tissue growth factor (CTGF/CCN-2), Hedgehog, Notch, and Wnt/β-catenin signaling. Regulatory pathways, including phosphoinositol-3 kinase (PI3K)/Akt, Hippo, peroxisome proliferator-activated receptors (PPARs), and farnesoid X receptor (FXR) (*9–11*), exhibit substantial variability across tissues. Despite the growing catalog of tissue-specific fibrosis markers and pathways, to the best of our knowledge, there are no established gene signatures or pathways that consistently capture fibrosis across various human organs. Identifying and validating these gene signatures or pathways and understanding how they integrate to drive the development and progression of fibrosis is essential for uncovering the underlying biology and guiding the development of effective, broadly applicable anti-fibrotic therapies.

In this study, we performed a comprehensive, cross-platform multi-layer transcriptomic analysis of human fibrosis across multiple organs, including the liver, kidney and heart, by integrating microarray, bulk RNA-seq, and single-nucleus RNA-seq datasets. By employing multi-center cohort and cross-platform data from a total of 1859 samples, and exploring different levels including single gene, pathway and network, we identified and verified a novel multi-organ fibrosis-associated gene signature (pan-fibrotic gene expression signature) that reflects conserved transcriptional alterations across organs. To our knowledge, this is the first study to identify and validate a pan-fibrotic signature conserved across the liver, kidney, and heart in humans using pan-transcriptomic data. Notably, our results suggested that fibrosis may be driven not only by excessive collagen production, but also by impaired ECM degradation. Our findings support this dual mechanism, highlighting the altered expression of genes involved in formation of collagen fibrils such as *COL5A1*, *ARHGEF2*, *SOX4*, *KLF15*, *CCL5*, *PLA2G4C*, *AZGP1*, alongside the downregulation of metallothioneins (MT; *MT1X* and *MT1E*), which are associated with reduced *MMP9* activation. Collectively, this study provides additional insights into molecular pathophysiology that may enable the discovery of conserved biomarkers applicable across multiple organ systems, improve early diagnosis, and support the design and development of broadly effective anti-fibrotic therapies.

## Results

### Identification of a conserved pan-fibrotic gene-expression signature in chronic diseases and its link to biology

Towards the discovery of a pan-fibrotic gene signature, we re-analysed multi-centric bulk microarray datasets comprising human tissue samples representing fibrotic and non-fibrotic conditions (n = 1051). Analysis at the level of each organ system enabled the identification of organ-specific fibrotic signatures, comprising 4523 differentially expressed genes in the heart (n = 555 samples), 1769 and 2391 genes in kidney tubuli (n = 144 samples) and glomeruli (n = 103 samples), respectively, and 1777 in the liver (n = 249 samples; see Supplementary Table S1-S3). By considering genes that were part of at least one organ-specific fibrotic signature and showed consistent directionality of regulation (when comparing fibrotic cases to controls in the discovery cohort), 572 genes were identified (Supplementary Table S9). To further validate their robustness, 325 additional samples, as available, from four bulk microarray datasets of liver tissues with known degrees of fibrosis were analyzed, confirming the regulation trend of 497 of them, resulting in the establishment of pan-fibrotic gene expression signature (Fig. 2a and Supplementary Table S10).

**Fig. 1:**
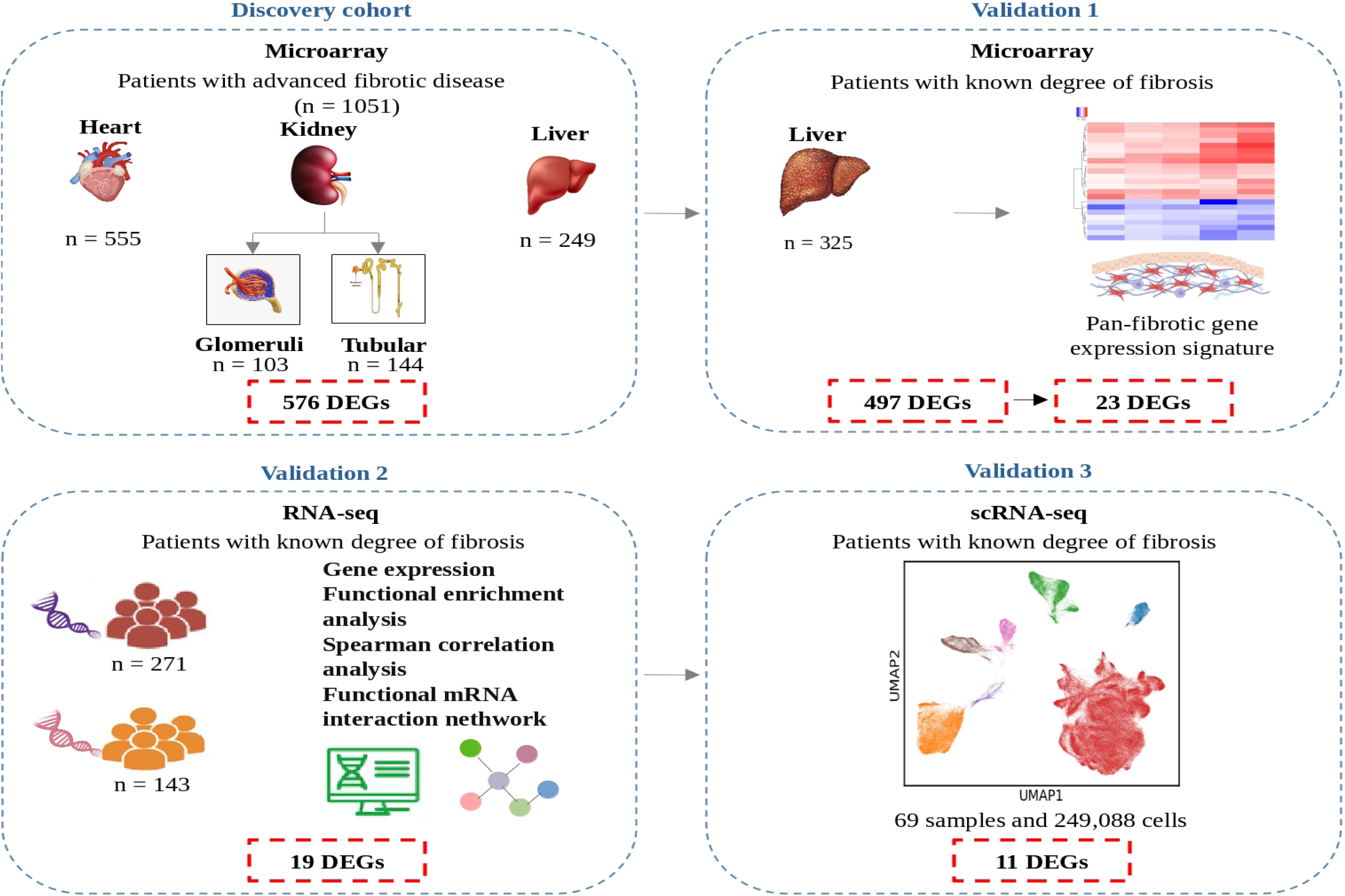
Schematic depiction of the study design for the discovery and validation of the pan-fibrotic gene expression signature. The analysis integrates multi-centric bulk microarray, RNA-seq, and single-cell RNA-seq (scRNA-seq) datasets to identify and validate shared fibrotic genes across organs. The number of pan-fibrotic genes identified at each stage is indicated in red squares with red dashed outlines. Abbreviations: DEGs, Differentially expressed genes; RNAseq, RNA sequencing; mRNA, messenger RNA; scRNAseq, single-cell RNA sequencing.

**Fig. 2:**
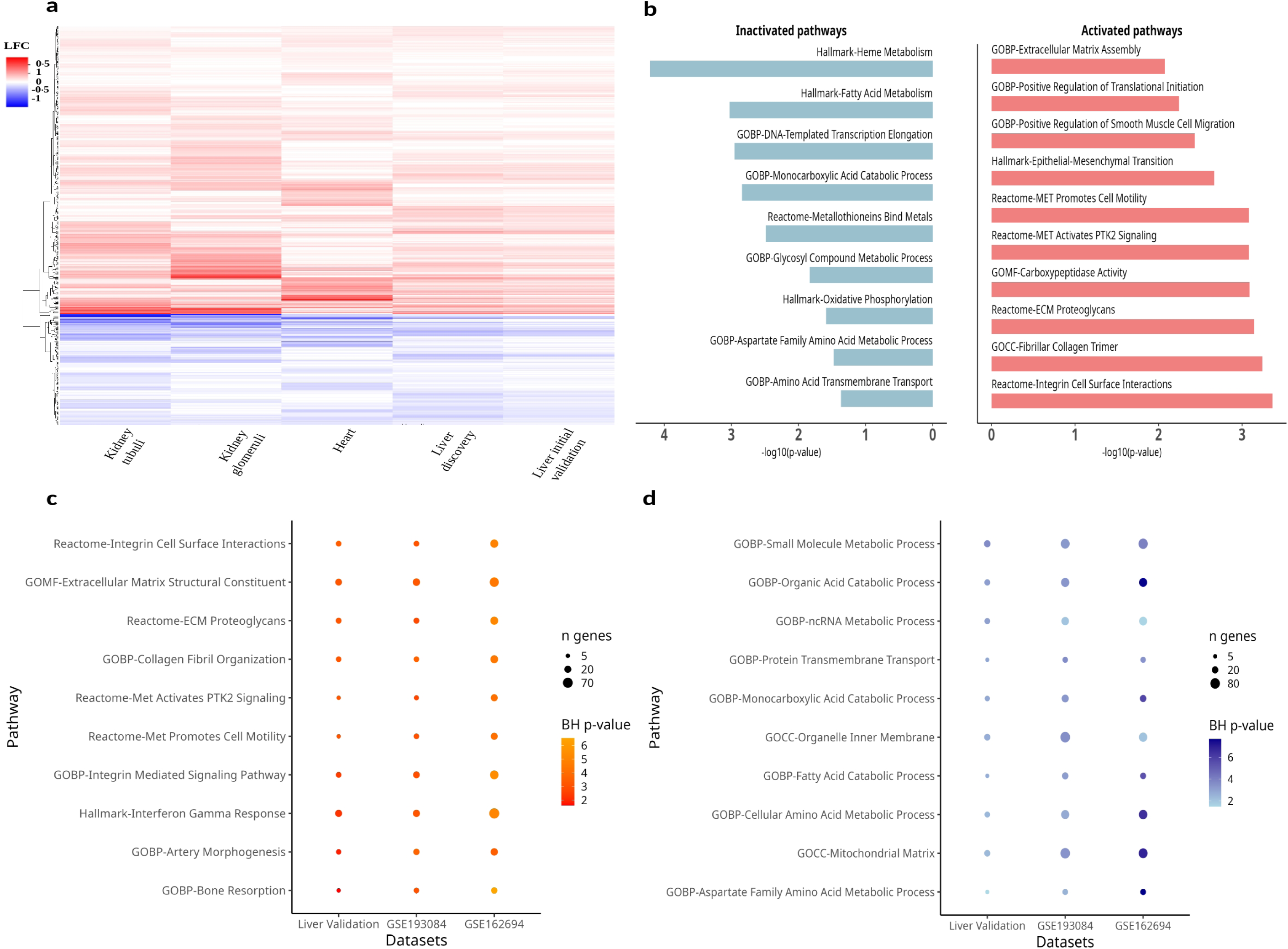
Identification and validation of a pan-fibrotic signature/pathways using multi-centric bulk microarray and RNAseq datasets. Heatmap representing the **(a)** 497 fibrotic signatures identified in the discovery and validation cohorts. Z-scaled fold changes are shown, with increased expression in fibrotic tissues depicted in red and decreased expression in blue. **(b)** Bar plot summarizing the top activated (red) and inactivated (blue) pathways across the discovery and initial validation cohorts, based on pathway enrichment analysis. **(c)** Dot plot showing the top activated pathways and (d) top inactivated pathways validated across the liver RNA-seq datasets (GSE193084 and GSE162694). Dot size represents the number of significant genes contributing to each pathway, while color indicates statistical significance (-log10(BH p-value)). The top enriched pathways were selected based on the lowest adjusted p-value in the microarray initial validation cohort and the highest coverage (defined as the proportion of genes contributing to each pathway). When multiple pathways were associated with the same biological process, the one with the highest coverage was retained. Abbreviations: GOBP, Gene Ontology-Biological Process; GOMF, Gene Ontology-Molecular Function; GOCC, Gene Ontology-Cellular Component; ECM, extracellular matrix; ncRNA, noncoding RNA.

To further investigate the molecular mechanisms underlying fibrosis, pathway activation analysis was conducted using the expression profiles of the 497 components of the pan-fibrotic gene expression signature in fibrotic versus non-fibrotic conditions across each affected organ system. The analysis revealed 172 activated and 143 inhibited pathways/processes that were significantly enriched across all organ systems in both the discovery and initial validation cohorts, all showing the same activation trend (Supplementary Table S11). The most significant activated molecular processes were predominantly enriched with pathways related to ECM remodelling. These included collagen fibril organization, ECM proteoglycans, and positive regulation of smooth muscle cell migration, highlighting the central role of ECM-related processes in promoting structural remodeling (Fig. 2b).

Conversely, inactivated molecular processes were primarily associated with metabolic pathways, including heme metabolism, fatty acid metabolism, and oxidative phosphorylation. Notably, zinc ion homeostasis pathways and cellular response to zinc ion, along with transcriptional regulation pathways, including DNA-templated transcription elongation, were significantly attenuated in fibrotic tissues (Fig. 2b; Supplementary Table S11). These findings suggest a disruption in cellular homeostasis, driven by metabolic dysregulation and impaired zinc ion signaling, which may contribute to the progression of fibrosis.

### Pathway-level validation across datasets

To validate the previously identified fibrosis-associated pathways/processes, gene set enrichment analysis was performed using the two independent RNA-seq datasets (n_RNAseq1_ = 271, n_RNAseq2_ = 143). Among the pathways that consistently maintained the same activation or inhibition status across all comparisons in the microarray analysis, 74 activated and 13 inhibited pathways were confirmed (BH p <0·05 and same trend in the fold change), representing the most conserved molecular processes associated with fibrosis across platforms (Supplementary Table S12).

Consistent with the microarray findings, the most significantly activated molecular processes in the RNA-seq datasets were associated with ECM remodelling and immune response. These included pathways such as collagen fibril organization, ECM proteoglycans, and integrin-mediated signaling. Immune-related processes, including interferon gamma response and activation of signaling pathways such as MET activates PTK2 signaling, were also among the top conserved activated pathways. This underscores the interplay between immune signaling and ECM remodeling in the pathophysiology of fibrosis (Fig. 2c; Supplementary Table S12).

Conversely, the verified inhibited pathways were primarily linked to metabolic processes, consistent with the findings from the microarray analysis. Notable examples included pathways associated with the mitochondrial matrix, such as oxidative phosphorylation and fatty acid metabolism, as well as transcriptional regulation processes, including DNA-templated transcription elongation. These inhibited pathways reflect a disruption in metabolic and transcriptional homeostasis, which may contribute to impaired cellular function and fibrotic progression (Fig. 2d; Supplementary Table S12).

### Hallmark elements of the pan-fibrotic gene-expression signature

To identify hallmark molecular features within the pan-fibrotic gene expression signature (497 genes), we assessed the data for genes that consistently showed statistically significant differential expression (BH p <0·05) across all comparisons in both discovery and initial validation sets (Fig. 3a; Supplementary Table S13). This resulted in the identification of 23 genes, among those well-establish genes implicated in fibrosis and ECM remodeling, such as *CCL5*, *SERPINE2*, *THBS2*, *COL5A1*, *VEGFC*, *SOX4*, among others, supporting the validity of the approach. Notably, the analysis also identified novel genes not previously implicated in fibrosis across organs, including *SYT11*, *CRIP1*, *PLA2G4C*, *ARHGEF2*, *CA2*, and *ELL2*, and members of the MT1 family (*MT1X* and *MT1E;* Fig. 3a).

**Fig. 3:**
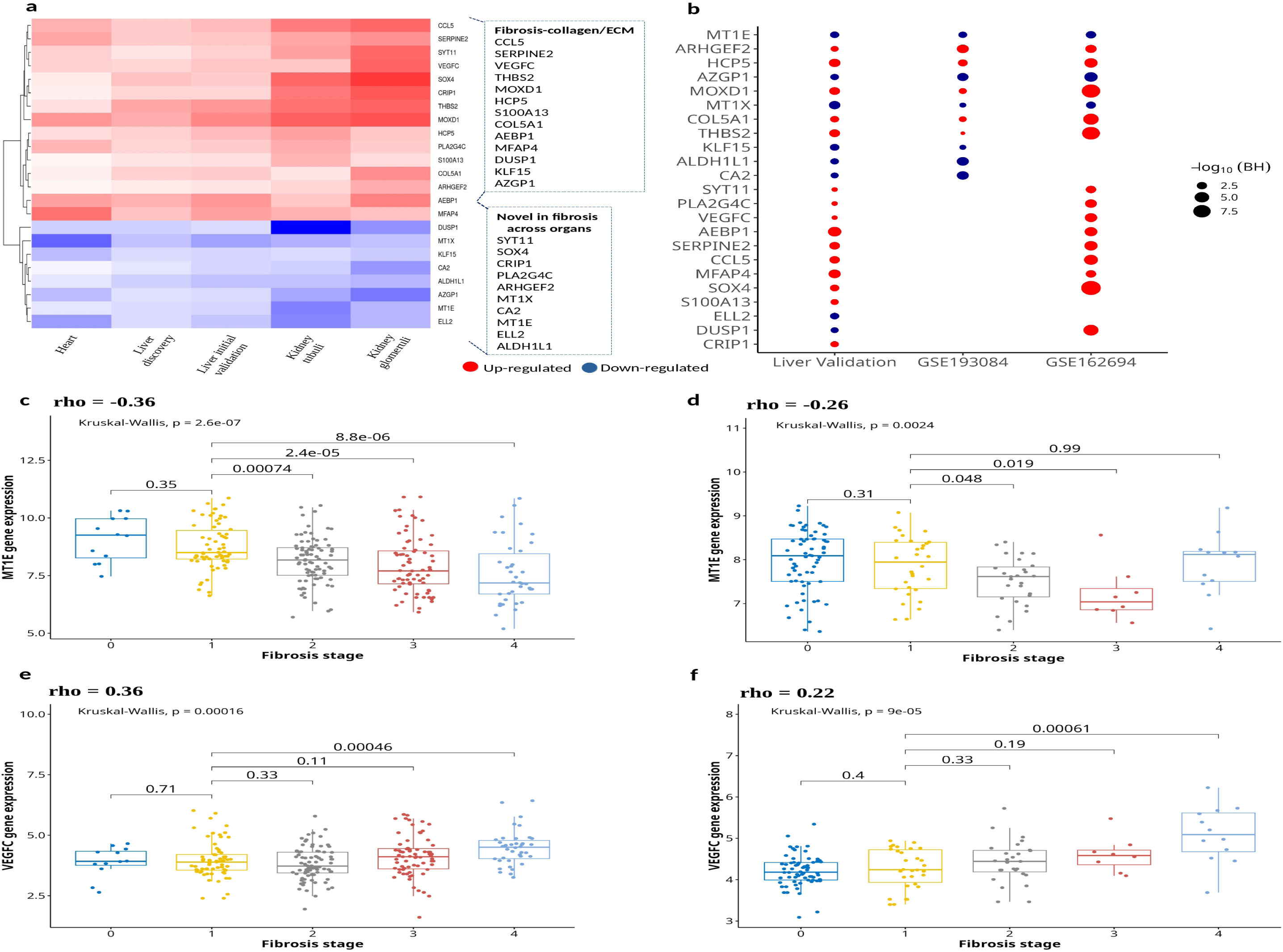
Hallmark elements of the pan-fibrotic gene-expression signature. **(a)** 23 hallmark molecular features within the pan-fibrotic gene expression signature. **(b)** Validation of the 23 fibrotic-associated genes in two independent RNA-seq liver datasets (GSE193084 and GSE162694). Each dot represents a significant gene (BH p <0·05), with genes showing increased expression in fibrosis represented in red and those with decreased expression in green. **(c)** Box plots illustrating the association of MT1E expression with fibrosis progression in the GSE193084, and **(d)** in the GSE162694 RNA-seq liver validation datasets. **(e)** Association of VEGFC expression with fibrosis progression in the GSE193084 and **(f)** in the GSE162694 RNA-seq liver validation datasets. Rho values indicate Spearman correlation coefficients, and p-values were calculated using the Kruskal-Wallis test. Abbreviations: ECM, Extracellular matrix.

To further verify the association of these 23 genes with fibrosis, we analyzed 414 samples from two independent RNA-Seq datasets, utilized as a second, cross-platform validation cohort (n_RNAseq1_ = 271, n_RNAseq2_ = 143; Supplementary Table S6). Nineteen genes were validated in at least one of the two RNA-seq datasets (GSE193084 and GSE162694), showing significant difference in expression (BH p <0·05) with consistent fold change orientation (Fig. 3b; Supplementary Table S13). Among these, six genes were downregulated (*MT1E, AZGP1, MT1X, KLF15, ALDH1L1,* and *CA2*), while thirteen were upregulated in the fibrotic conditions (*ARHGEF2, HCP5, MOXD1, COL5A1, THBS2, SYT11, PLA2G4C, VEGFC, AEBP1, SERPINE2, CCL5, MFAP4,* and *SOX4*). Further investigation of association of gene expression with the degree of fibrosis revealed significant correlation for 15 of the 19 validated genes in both RNA-seq datasets (BH p <0·05). These genes exhibited consistent positive (*ARHGEF2, HCP5, MOXD1, COL5A1, THBS2, SYT11, PLA2G4C, VEGFC, AEBP1, CCL5, MFAP4,* and *SOX4)* or negative (*MT1E, AZGP1,* and *MT1X)* correlations with fibrosis severity across the two RNAseq datasets, reinforcing their potential as conserved biomarkers or regulators of fibrotic progression (Fig. 3c-f; Supplementary Fig. S7a-h; Supplementary Table S14).

To achieve cell-level resolution in our analysis, we investigated the cell-type-specific expression patterns of these hallmark genes using an integrated human multi-cohort snRNA-seq liver dataset (Fig. 4a). Cell types were identified based on canonical liver cell markers, enabling reliable annotation of major hepatic populations within the integrated dataset (Fig. 4b). A chi-square of independence revealed a highly significant association between cell type distribution and fibrosis severity (BH p < 0·0001). The proportion of hepatic stellate cells (HSC), T cells, LSCEs, hepatocytes, and endothelial cells populations showed significant changes, particularly in moderate and severe fibrosis, suggesting their active involvement in fibrosis progression (Fig. 4c). We assessed the contribution of individual cell types to the expression of the 19 validated genes and found that all major hepatic cell populations contributed to their expression (Fig. 4d). To further shortlist the pan-fibrotic gene expression signature, differential expression analysis identified 11 out of the 19 genes as statistically significant, with consistent fold change directionality between fibrotic and non-fibrotic conditions, as previously observed (Fig. 4e). The shortlisted features included *KLF15*, *ALDH1L1*, *PLA2G4C*, *VEGFC*, *CLL5*, *SOX4*, *MT1E*, *ARHGEF2*, *AZGP1*, *MT1X*, and *COL5A1* (Supplementary Table S15). Genes directly or indirectly involved in ECM production and deposition (e.g., *SOX4*, *ARHGEF2*, *VEGFC*, *CCL5*, and *COL5A1*) exhibited distinct, cell-type-specific expression profiles; while MT family members, involved in zinc ion homeostasis (e.g., *MT1X*, *MT1E*), were predominantly expressed in hepatocytes and liver sinusoidal endothelial cells (LSECs). Interestingly, some genes were specifically differentially expressed in certain cell types (e.g. *PLA2G4C in* hepatic stellate cells (HSCs) and endothelial cells), suggesting a potential cell-type-specific regulatory role in fibrotic progression. At the same time, other genes (e.g. *AZGP1)* were significantly downregulated across all major liver cell populations, suggesting a pan-cellular fibrotic response.

**Fig. 4:**
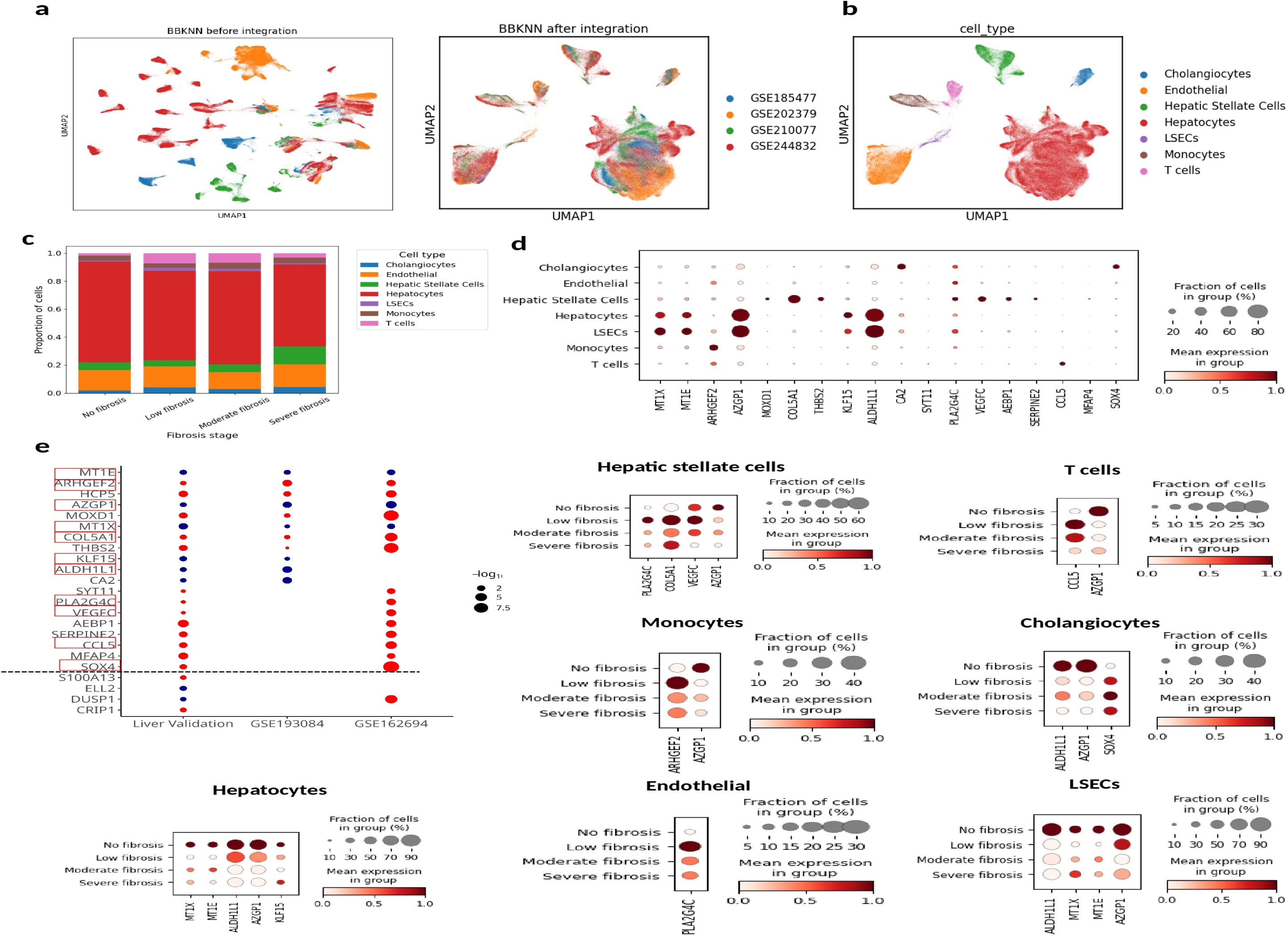
Single-cell validation of the pan-fibrotic gene expression signature in an integrated multi-cohort human liver snRNA-seq dataset. **(a)** UMAP plots showing the unintegrated (left panel) and integrated (right panel) snRNA-seq datasets from four independent cohorts (total cells = 249,088). **(b)** UMAP displaying the annotated liver cell types identified following integration. **(c)** Proportional distribution of liver cell types across fibrosis stages in the integrated multi-cohort snRNA-seq dataset. **(d)** Dot plot showing cell-type–specific expression patterns of the 19 validated hallmark pan-fibrotic genes. **(e)** Validated genes using snRNA-seq are highlighted in red (top left). Dot plots illustrating the expression of the 11 single-cell validated hallmark pan-fibrotic genes across cell types and fibrosis severity. Abbreviations: BBKNN, Batch Balance K-Nearest Neighbors; UMAP, Uniform Manifold Approximation and Projection; LSECs, liver sinusoidal endothelial cells.

### Network-level analysis reveals functional connectivity of key components

To elucidate the functional roles and molecular interactions of the 11 hallmark pan-fibrotic genes, we constructed a PPI network. This analysis revealed a high degree of interconnectivity among the 11 hallmark pan-fibrotic genes and other molecules implicated in key biological pathways. The network clustered into distinct modules enriched for biological processes such as ECM organization, signal transduction, immune response, metabolism, regulation of transcription, and metal ion homeostasis (Fig. 5).

**Fig. 5:**
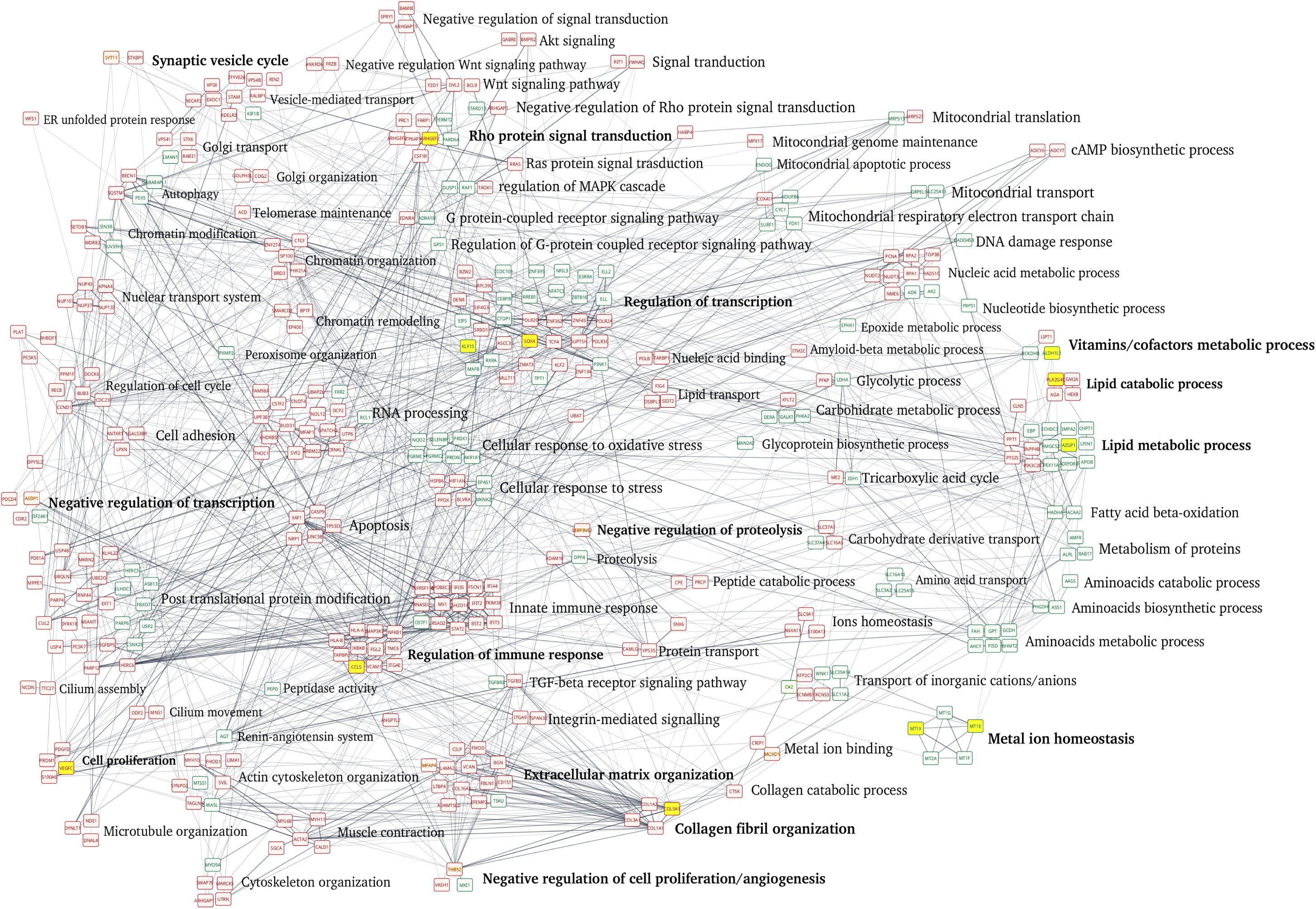
Functional PPI analysis of fibrosis-associated genes. 11 validated hallmark pan-fibrotic genes are highlighted in yellow. Upregulated differently expressed genes are depicted in red, while down-regulated genes are depicted in green. Abbreviations: ER, endoplasmic reticulum; Rho, Rho family small GTPase; Ras, Ras family small GTPase; MAPK, mitogen-activated protein kinase; cAMP, cyclic adenosine monophosphate; TGF, transforming growth factor.

In this study, *MT1X* and *MT1E* were identified as novel potential cross-organ fibrosis markers. Within the network, *MT2A* and *MT1E*, central components of the “metal ion homeostasis” module, were found to interact with several genes across three clusters, *STAT2*, *SLC39A14*, *PISD*, and *BHMT2.* These connections suggest a mechanistic link between metal ion homeostasis, transport of inorganic cations/anions, cellular metabolism, and immune regulation. Moreover, indirect interactions mediated through *CRIP1*, between *CA2* and *CTSK*, suggests a potential association between transport of inorganic cations/anions and collagen catabolic processes in fibrotic pathology. Notably, the hallmark pan-fibrotic genes were identified in various biological processes within the network (Fig. 5). These genes mediate interactions across diverse fibrosis-associated pathways, highlighting the complexity and functional interconnectivity underlying fibrotic pathogenesis. Collectively, our findings suggest that coordinated dysregulation of these interconnected pathways may drive fibrotic progression and highlight them as potential targets for therapeutic intervention.

## Discussion

Fibrosis is a common pathological endpoint across numerous chronic diseases and a major contributor to progressive organ dysfunction and failure (*2*). Despite extensive research into tissue-specific fibrotic mechanisms, few studies have systematically investigated whether shared molecular signatures/pathways exist across different organs (*12*). This knowledge gap limits the discovery of broadly applicable pan-biomarkers and anti-fibrotic therapies, which could be particularly beneficial in chronic disease of heart, liver and kidney. In this context, our study aimed to identify a common fibrotic signature through a rigorous, large-scale, multi-cohort, multi-tissue and multi-level transcriptomic analysis (bulk microarray, RNA-seq, and single-cell RNA-seq data) covering a total of 1859 samples. Through this integrative approach, we defined a pan-fibrotic gene expression signature comprising of 497 genes consistently dysregulated in fibrotic tissues. Among those, 11 were shortlisted as hallmark features, being validated across all data types (Fig. 6; Supplementary Table S15). Notably, several of these hallmark genes including *MT1E*, *MT1X*, *ARHGEF2*, *SOX4*, *PLA2G4C,* and *ALDH1L1*, have not previously been implicated in fibrosis in humans, shedding additional insights into conserved pathophysiological mechanisms of fibrosis.

**Fig. 6:**
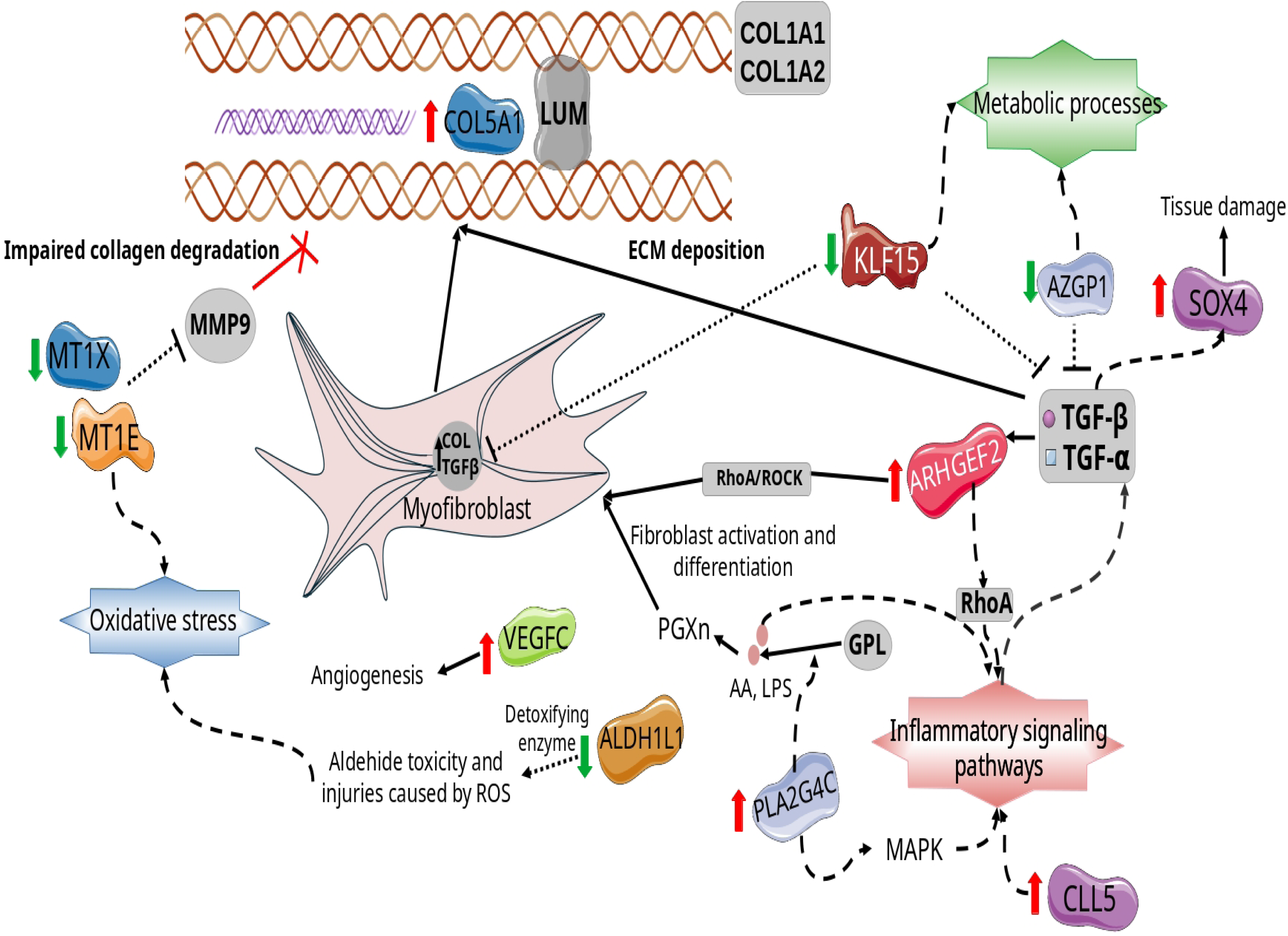
Schematic summary of the hallmark pan-fibrotic genes and their potential functional relevance to fibrosis. The 11 hallmark pan-fibrotic genes genes identified in this study are shown in color, while previously known fibrosis-related components (e.g., TGF-β, α-SMA, RhoA/ROCK) are shown in grey for contrast. Abbreviations: GPL, glycoprotein ligand; AA, arachidonic acid; LPS, lipopolysaccharide; PGXn, prostaglandins; ROS, reactive oxygen species; RhoA/ROCK, Ras homolog family member A/Rho-associated protein kinase; COL, collagen.

Previous studies have demonstrated that the development of fibrosis is, in part, driven by excessive collagen production (*2*, *12*, *13*). In line with this, our analysis identified altered expression of several fibrillogenic genes, including *COL5A1, ARHGEF2, SOX4, KLF15, CCL5, PLA2G4C,* and *AZGP1*. Among the novel genes, *ARHGEF2* was notably upregulated in fibrotic tissues, suggesting a potential role as a key mediator of fibrotic progression. *ARHGEF2* is a guanine nucleotide exchange factor that activates Rho GTPases, particularly RhoA, involved in ECM production, activation of immune response, and cytoskeletal remodeling (*14*). In vitro studies have shown that silencing *ARHGEF2* reduces the expression of pro-fibrotic markers such as α-smooth muscle actin (*α-SMA*) and prevents epithelial-to-mesenchymal transition (EMT) in kidney and retinal cells (*14*, *15*). Similarly, in vivo murine models of cardiac and renal fibrosis have demonstrated elevated *ARHGEF2* expression, while its inhibition attenuates fibrosis, cardiomyocyte apoptosis and the development of chronic heart failure (*16*). Given upstream regulatory role in multiple pro-fibrotic pathways, *ARHGEF2* may represent a promising therapeutic target for modulating fibrotic responses across tissues.

In line with this, the transcription factor *SOX4* was upregulated in fibrotic tissues. *SOX4* belongs to the SOX family of transcription factors, which play critical roles in tissue morphogenesis, development, and various disease processes (*17*). Although *SOX4* has not been extensively studied in the context of fibrosis across organs, emerging evidence suggests a potential role. In cardiac pathology, *SOX4* expression correlates with fibrotic gene signatures in both dilated and hypertrophic cardiomyopathy, suggesting its involvement in this process (*18*). Furthermore, in a liver injury model, *SOX4* expression was induced by injury-associated signals such as TGF-β, Notch, YAP, contributing to pathological remodeling and tissue damage (*17*). Interestingly, this regulatory response parallels the dynamics observed in *SOX9*-driven regeneration, where sustained activation determines whether tissue regeneration proceeds adaptively or results in fibrosis (*19*).

Moreover, our study identified upregulation of *PLA2G4C*, a member of the phospholipase A2 (PLA2) superfamily of lipolytic enzymes. *PLA2G4C* catalyzes the hydrolysis of glycerophospholipids, leading to the release of free fatty acids such as arachidonic acid and lysophospholipids. This reaction represents a key step in the biosynthesis of pro-inflammatory prostaglandins, which are known to influence fibroblast activation and differentiation, thereby contributing to fibrotic remodeling (*20*). Furthermore, different secretory *PLA2* isoforms were found upregulated in fibroblast of patients with Idiopathic Pulmonary Fibrosis (IPF) and treatment with a known sPLA2 inhibitor attenuated lung fibrosis along with a reduction in *TGF-β* and deposition of ECM in lung (*21*). Beyond gene-level findings, our study reveals key dysregulated biological processes that are conserved across fibrotic organs. Among the activated pathways, those related to ECM remodeling and integrin signaling, as well as previously described core signaling pathways (*2*, *8*, *9*), were predominant. This finding underscores the central role of structural reorganization in fibrosis (Supplementary Table S12).

Interestingly, we identified *MT1E* and *MT1X*, members of the MT family, as significantly downregulated across all tissues and negatively correlated with fibrosis severity, together with pathways related to MT-bind metals, zinc ion homeostasis and cellular response to zinc ion pathways (Supplementary Table S12; Supplementary Table S15). These findings suggest that impaired intracellular zinc homeostasis may represent an underexplored, common tissue mechanism in fibrosis biology. Notably, the primary transmembrane zinc transporter *SLC39A14* was also downregulated across all tissues (Supplementary Table S11). Given that *SLC39A14* mediates cellular zinc uptake, its reduced expression could contribute to the suppression of MT via reduced intracellular zinc availability (*22*, *23*). Other MT isoforms (*MT2A*, *MT1F,* and *MT1G)* showed similar trend of downregulation, although not reaching statistical significance across all tissues (BH <0·05), further supporting the findings. To our knowledge, this is the first report of consistent downregulation of MTs and *SLC39A14* in association with fibrotic progression across multiple human organs, highlighting their potential as novel, cross-organ fibrosis markers. This finding is consistent with pre-clinical and clinical evidence linking zinc imbalance to fibrosis affecting the heart, kidney and liver (*24–27*), as well as findings in *Slc39a14* knockout mice, which exhibit reduced zinc levels and lower *Mt-1* expression (*28*). Notably, in murine liver fibrosis models, zinc supplementation restored *Slc39a14* expression and reverse fibrotic process (*29*), reinforcing the role of these genes as mediators of zinc homeostasis in fibrosis progression.

Although direct links between *MT1E* and *MT1X* isoforms and fibrosis or ECM remodeling are limited, prior studies in cell lines have shown that silencing of related isoform *MT2A* in cell lines reduces *MMP9* activity (*30*, *31*). This suggests that *MT1E* and *MT1X* downregulation, together with reduced zinc ion availability, may impair *MMP9* catalytic function and limit ECM degradation (*27*, *30–32*). Notably, in our study several *MMPs* showed increased expression in fibrotic tissues, likely reflecting a compensatory response to support matrix degradation. However, in part by a zinc-deficient environment, their enzymatic activity may be compromised. Concurrent upregulation of *TIMP1* (as indicated by our results) in this environment may further inhibit *MMP* function, thereby in part contributing to impaired resolution of fibrosis (Supplementary Table S11) (*29*). These findings are consistent with previous in vivo studies. MT knockout mice developed increased aortic fibrosis in a mouse model of intermittent hypoxia (*33*). Conversely, MT-overexpressing transgenic (MT-TG) mice demonstrated protection against progressive myocardial fibrosis during the long term follow-up (1 to 6 months) and against liver fibrosis (*34*, *35*). Taken together, our findings suggest a potential anti-fibrotic role for *MT1E* and *MT1X*, likely through their involvement in regulating *MMP*-dependent ECM degradation. While fibrillogenic genes are broadly expressed across tissues and contribute to excessive collagen production, the consistent downregulation of MT and zinc homeostasis pathways points to an additional mechanism, complementary mechanism: impaired ECM degradation. Together, these data support the notion that fibrosis is driven not only by increased ECM synthesis, but also by the failure to adequately degrade and remodel the matrix.

Furthermore, additional consistently inactivated processes involved mitochondrial metabolism and transcriptional regulation. Metabolic pathways were consistently found to be inactivated across fibrotic organs, including organic acid catabolic process, monocarboxylic acid catabolic process, fatty acid catabolic process, and cellular amino acid catabolic process (Gene Ontology: Biological Process, GOBP). These findings are in line with recent evidence that associated metabolic reprogramming with chronic diseases, where changes in cellular metabolism have been shown to promote myofibroblast activation and improve collagen stabilization (*36*, *37*). Overall, our results indicate that metabolic dysfunction, together with disturbed metal ion regulation, represents a complementary layer of dysregulation that may shape the fibrotic phenotype across tissues.

Moreover, snRNAseq analysis revealed that the proportion and transcriptional activity of cell populations shifted across fibrosis stages, revealing a more complex cellular architecture than previously described. Although in previous studies HSCs are well-established as key mediators of fibrosis due to their central role in ECM production (*38*, *39*), our findings indicate that other hepatic cell types actively contribute to fibrosis progression. Together, our results offer new insights on the cell-type-resolved regulation of fibrosis-associated gene signatures in human liver tissue.

The PPI network analysis further demonstrated that several hallmark pan-fibrotic genes act as key regulatory nodes, linking different fibrotic processes including ECM organization, immune signaling, metabolic activity, and metal ion homeostasis. These findings reflect the complexity nature of fibrosis, indicating that its progression involves the coordinated disruption of multiple, interdependent biological systems rather than isolated pathways. Importantly, some pan-fibrotic genes not previously associated with fibrosis, such as *MT1E*, *MT1X*, *ARHGEF2*, *SOX4*, *PLA2G4C,* and *ALDH1L1,* were observed across distinct pathways, suggesting possible roles in mediating cross-pathway communication during fibrotic remodeling.

Collectively, by integrating multi-organ transcriptomic data from the heart, liver, and kidney, this study defined a pan-fibrotic gene signature and shared regulatory pathways, highlighting potential new regulators of fibrosis, thus providing additional insights into the conserved molecular mechanisms underlying this pathological process. These insights underscore the need for multi-targeted therapeutic approaches and lay the groundwork for cross-organ, precision medicine strategies in fibrosis treatment.

### Limitations and strengths

Our study has several limitations arising from the analysis of publicly available data. Firstly, the discovery cohort included patients with chronic diseases known to involve fibrosis, but the detailed metadata on fibrosis was not available. However, a major strength addressing this limitation is the rigorous validation strategy: findings from the discovery cohort were confirmed across multiple platforms and independent cohorts with comprehensive fibrosis-stage annotation, significantly strengthening reproducibility and robustness. Secondly, the present study is not supported by direct experimental validation in animal models or cell lines, which would provide mechanistic insight into the identified genes. Nevertheless, the robustness and translational relevance of our findings are reinforced by the large-scale, multi-organ integration analysis of thousands of human samples across multiple datasets and tissues. Additionally, existing literature from animal and cell-line models, as highlighted in our discussion, supports our findings and underscores the therapeutic potential of the identified genes, thus paving the way for future pre-clinical and mechanistic studies.

## Materials and Methods

### Study design and data retrieval

To investigate the conserved molecular fibrotic signature across various organs, publicly available transcriptomics data acquired from the liver, kidney, and heart tissues, obtained from patients diagnosed with chronic disease involving fibrosis, were retrieved from Gene Expression Omnibus, and analyzed through a multi-step and cross-platform process, as depicted in Fig. 1. Details on the systematic data retrieval and curation of both publicly available microarray, RNA-seq, and scRNA-seq data are presented in Supplementary Fig. S1; Supplementary Methods.

A total of 1236 microarray datasets from human tissue samples (specifically from heart, kidney and liver) were retrieved. Due to limited availability of information about the stage of fibrosis, microarray datasets were divided into 2 groups: a discovery cohort comprising studies of chronic heart, kidney and liver diseases known to be associated with fibrosis but without fibrosis stage information available (16 studies; n=1051; Supplementary Table S1-S3) and validation cohort with available staging data (4 studies; n = 325; Supplementary Table S4). Moreover, 414 liver tissue samples from two RNAseq datasets with detailed fibrosis stage information were retained and included as a second independent validation cohort (Supplementary Table S5).

Finally, four publicly available single-nuclei RNA-seq (snRNA-seq) datasets of human liver tissue retrieved from the GEO repository were also included as part of the validation process (Supplementary Table S6). In this case, the analysis focused specifically on liver fibrosis, as these datasets included samples with annotated fibrosis stages determined by histological assessment of routine staining, as described previously in (*40–42*). In total, datasets from 69 liver samples covering different fibrosis stages were re-analyzed. Given the heterogeneity in fibrosis staging systems across studies, liver fibrosis scores were harmonized into low, moderate, and severe categories based on standardized histological classifications (Supplementary Table S7) (*43–47*).

### Bulk transcriptomic dataset retrieval and processing

All raw microarray data were uniformly retrieved, processed, and then integrated within each organ system, following the previously described pipeline (*48*). To ensure comparability across the different GEO datasets within the organ system, batch effect removal was applied to gene expression data using the ComBat algorithm (*49*). This adjustment was evaluated using relative log expression and principal component analysis plots, supporting the successful removal of the batch effect (Supplementary Fig. S2-S5). Importantly, biological variability was preserved, as evidenced by the sustained high expression levels of heart-specific housekeeping genes such as *CASQ2*, *CRYAB*, *UBB* and *MYL3* in the merged heart tissue dataset (Supplementary Fig. S6). These results indicate that application of ComBat successfully removed batch effects while preserving biological variability.

For the analysis of bulk RNAseq data, the raw FASTQ files were aligned to the human reference genome sequence (hg38) using STAR with default parameters, and the package Rsubread (v1.5) was used to generate summary counts. RNA-seq data were analyzed per dataset to further increase confidence in the reproducibility of the findings. Detailed information about the bulk transcriptomics data processing is presented in the Supplementary Methods.

### snRNAseq data processing

Data processing and downstream analysis were performed using the Python package Scanpy (v1.11.0). Raw count matrices were obtained and loaded into Scanpy objects for quality control filtering. Cells expressing fewer than 500 or more than 5,000 genes, or exhibiting over 10% mitochondrial transcript content, were excluded. After quality filtering, the resulting AnnData objects from individual samples were merged into a single dataset for subsequent integration and analysis. A total of 249,088 high-quality cells with 17,684 genes were retained. To perform dimensionality reduction, batch correction, and clustering, the top 4,000 highly variable genes across batches were identified. The batch effect arising from diverse platforms and protocols was subsequently removed with the BBKNN algorithm using the Python package BBKNN (v1.6.0). Cell clusters were manually annotated according to the expression of canonical marker genes for each cluster (Supplementary Table S8). Further details are provided in the Supplementary Methods.

### Statistical analysis and defining the pan-fibrotic gene expression signature

Statistical analyses of the bulk transcriptomics data was performed using the R software (v4.1.0), including the stats package for the Wilcoxon rank-sum test (for microarray), the DESeq2 package for the negative binomial Wald test (for RNA-seq), and the stats package for the Spearman’s correlation. False discovery rate (FDR) correction was applied using the Benjamini-Hochberg (BH) method, and adjusted p < 0·05 was considered statistically significant. In all cases, comparisons were made between “fibrotic” and “non-fibrotic” conditions.

In the snRNAseq data, cell proportions differences across fibrotic stages were compared with the chi-square test. Differentially expressed genes between the ‘fibrosis’ and ‘no fibrosis’ groups were identified within each cluster using the Python package Scanpy (v1.11.0), “rank_genes_groups” function, and the Wilcoxon rank-sum test. Differentially expressed genes were defined on BH p <0·05 and a minimum of 20% of cells expressing the gene in the higher-expression group.

To define pan-fibrotic gene expression signature based on the discovery microarray data, statistical analysis was first conducted with each organ system based on the integrated combat corrected data. To ensure that the findings can be also reproduced in the individual studies (without applying batch correction), genes found significant and with consistent fold change direction in individual datasets compared to the integrated data within each organ, were only considered, resulting in the establishment of organ-specific fibrotic signatures. The pan-fibrotic gene expression signature was defined as genes significantly differentially expressed in at least one fibrosis-affected organ system (BH p <0·05), while consistently maintaining the same fold change directionality across the different organs; in addition, the directionality of the fold change of each signature genes was further verified based on the microarray-derived dataset with known information about the fibrosis stage.

To further shortlist the key components of the pan-fibrotic gene expression signature, significance across all organ systems was required. This was followed by validation of both the direction of regulation and statistical significance in two bulk RNA-seq datasets and an integrated snRNA-seq dataset.

### Pathway enrichment analysis

Pathway activation scores in the bulk transcriptomics data were calculated using the pan-fibrotic gene expression signature (497 genes) with the “ssGSEA-GSVA” GSVA function. The analysis was performed separately for each organ system, based on the combat corrected integrated microarray discovery, as well as on the RNA-seq validation datasets. Pathway analysis of the RNA-seq data was conducted separately for each dataset using all respective significantly affected genes identified per dataset (BH p <0·05). Gene libraries from Hallmark (v7.5.1), Canonical Pathways (Reactome subset, v7.5.1), Gene Ontology (GO, BP, CC, MF v7.5.1), and Transcription Factor Targets (GTRD, v7.5.1), were retrieved from the Molecular Signature Database (MSigDB) using the R software (v4.1.0). Pathway concordance was defined as consistent enrichment (BH p <0·05) with the same direction of activation status (activation or inactivation) across all fibrosis-affected organ systems in the discovery and initial validation cohort. Subsequently, these fibrosis-associated pathways/processes were validated using the RNA-seq datasets. Those pathways found to be significantly enriched (BH p <0·05) in each RNA-seq dataset, and maintained the same activation or inhibition status in the microarray comparisons were considered as validated.

### Functional protein-protein interaction network

In order to explore the functional connectivity and potential regulatory hubs among the hallmarks of the pan-fibrotic gene expression signature (11 genes), a protein-protein interaction network (PPI) was generated using the STRING database (v2.2.0) and visualized with Cytoscape software (v3.10.3) (*50*), with interaction scores set at a medium confidence threshold (≥0·4). The analysis was based on 11 pan-fibrotic genes and other components within the activated and deactivated pathways identified in the discovery and initial validation cohort in which these shortlisted genes were involved. Moreover, to assess the functional context of genes within the PPI network, a manual assignment based on a literature review of biological processes associated with each gene was performed. Genes were grouped into clusters that reflected shared biological processes.

## Ethics

All datasets used in this study were publicly available and de-identified. Ethical approval for the use of human data was obtained by the original study investigators.

## Code availability

Code to reproduce the analyses described in this manuscript can be accessed via: https://zenodo.org/records/16570795.

## Acknowledgments

MAJC holds a doctoral grant through the DisCo-I project that has received funding from the European Union’s Horizon Europe Marie Skłodowska-Curie Actions Doctoral Networks - Industrial Doctorates Programme (HORIZON – MSCA – 2021 – DN-ID) under grant agreement No 101072828. We gratefully acknowledge the help of Dr. Jason Iacovoni for training Mayra Alejandra Jaimes Campos in single transcriptome analysis and normalization.

## Contributors

MAJC, HM, AL, AV, and JPS designed the study. MAJC conducted the collection and analysis of the data, generated the tables, figures, and wrote the original draft of the manuscript. RS contributed with the data analysis. RS and AL accessed and verified the data. MAJC, JCJ, MB, and AL contributed with the data interpretation. MAJC, HM, AL, AV, RS, JCJ, MB, and JPS review and editing the draft of the manuscript. All authors read and approved the final manuscript.

## Data sharing statement

Public data used in this work are available from the GEO database. The specific data accession numbers are listed in Supplementary Table S1-S7. All data generated during this study are included in the manuscript and supporting files. The statistical results from the transcriptomics analysis in the different tissues, along with all analysis scripts are available on https://zenodo.org/records/16570795.

## Supplementary data

### Supplementary tables

**Supplementary Table S1. Selected heart tissue microarray datasets.**

**Supplementary Table S2. Selected kidney tissue microarray datasets from tubules and glomeruli.**

**Supplementary Table S3. Selected liver tissue microarray datasets.**

**Supplementary Table S4. Selected microarray initial validation cohort datasets.**

**Supplementary Table S5. Independent RNA-seq second validation cohort.**

**Supplementary Table S6. Independent snRNA-seq validation cohort.**

**Supplementary Table S7. Unification of the different systems describing the degree of liver fibrosis in the microarray validation cohort.**

**Supplementary Table S8. Canonical markers for cell cluster annotation.**

**Supplementary Table S9. List of 572 validated pan-fibrotic genes differentially expressed between fibrotic and non-fibrotic samples, identified through integrated analysis of microarray datasets.**

p-values were adjusted using the Benjamini–Hochberg (BH) method. Significant changes (p<0·05) are marked in yellow. Fold changes labelled in red represent increased gene expression in fibrosis, and those in green represent decreased gene expression.

**Supplementary Table S10. List of 497 validated pan-fibrotic genes differentially expressed between fibrotic and non-fibrotic samples, identified through integrated analysis of microarray datasets.** p-values were adjusted using the Benjamini–Hochberg (BH) method. Significant changes (p<0·05) are marked in yellow. Fold changes labelled in red represent increased gene expression in fibrosis, and those in green represent decreased gene expression. In bold are the 23 MFGSig that consistently demonstrated significant changes (BH p-value <0·05) across all comparisons.

**Supplementary Table S11.Pathway analysis between fibrosis and non-fibrosis groups based on the analysis of microarray datasets.** p-values were adjusted using the Benjamini–Hochberg (BH) method. Significant changes (p<0·05) are marked in yellow. Fold changes labelled in red represent increased gene expression in fibrosis, and those in green represent decreased gene expression.

**Supplementary Table S12. Pathway analysis between fibrosis and non-fibrosis groups based on the analysis of microarray and RNA-seq datasets.** p-values were adjusted using the Benjamini– Hochberg (BH) method. Significant changes (p<0·05) are marked in yellow. Fold changes labelled in red represent increased gene expression in fibrosis, and those in green represent decreased gene expression.

**Supplementary Table S13. List of 23 validated hallmark molecular features differentially expressed between fibrotic and non-fibrotic samples, identified through integrated analysis of microarray and RNA-seq datasets.** p-values were adjusted using the Benjamini–Hochberg (BH) method. Significant changes (p<0·05) are marked in yellow. Fold changes labelled in red represent increased gene expression in fibrosis, and those in green represent decreased gene expression.

**Supplementary Table S14. Correlation of 11 validated hallmark molecular features with the degree of fibrosis in the snRNA-seq datasets.** p-values were adjusted using the Benjamini–Hochberg (BH) method. Significant changes (p<0·05) are marked in yellow. Rho values labelled in red represent increased gene expression in fibrosis stages, and those in green represent decreased gene expression in fibrosis stages.

**Supplementary Table S15. List of 11 validated hallmark molecular features differentially expressed between fibrotic and non-fibrotic samples, identified through integrated analysis of snRNA-seq datasets.** p-values were adjusted using the Benjamini–Hochberg (BH) method. Significant changes (p<0·05) are marked in yellow. Fold changes labelled in red represent increased gene expression in fibrosis, and those in green represent decreased gene expression.

### Supplementary figures

**Supplementary Fig. S1. Overall workflow for the retrieval and curation of microarray, and RNAseq publicly available data representing fibrotic and non-fibrotic conditions.** Abbreviations: RNAseq, RNA sequencing.

**Supplementary Fig. S2. Batch effect correction of heart microarray datasets (discovery). a-b.** Box plots and PCA plots of unadjusted expression data, showing dataset-specific variability. **C-D.** Box plots and PCA plots after ComBat adjustment, illustrating successful batch effect correction with improved sample distribution and reduced dataset-specific clustering. Abbreviations: PC, Principal Component.

**Supplementary Fig. S3. Batch effect correction of kidney tubuli microarray datasets (discovery). a-b.** Box plots and PCA plots of unadjusted expression data, showing dataset-specific variability. **c-d.** Box plots and PCA plots after ComBat adjustment, illustrating successful batch effect correction with improved sample distribution and reduced dataset-specific clustering. Abbreviations: PC, Principal Component.

**Supplementary Fig. S4. Batch effect correction of kidney glomeruli microarray datasets (discovery). a-b.** Box plots and PCA plots of unadjusted expression data, showing dataset-specific variability. **c-d.** Box plots and PCA plots after ComBat adjustment, illustrating successful batch effect correction with improved sample distribution and reduced dataset-specific clustering. Abbreviations: PC, Principal Component.

**Supplementary Fig. S5. Batch effect correction of liver microarray datasets (discovery + initial validation cohorts). a-b.** Box plots and PCA plots of unadjusted expression data, showing dataset-specific variability. **c-d.** Box plots and PCA plots after ComBat adjustment, illustrating successful batch effect correction with improved sample distribution and reduced dataset-specific clustering. Abbreviations: PC, Principal Component.

**Supplementary Fig. S6. ComBat-adjusted expression of housekeeping genes across individual heart microarray datasets. a-d.** Box plots showing the high and consistent mean expression levels of selected housekeeping genes after batch effect correction using the ComBat algorithm.

**Supplementary Fig. S7. Box plots illustrating the association of hallmark elements of the pan-fibrotic gene-expression signature, with fibrosis progression in the RNA-seq liver validation datasets. a-h.** Box plots showing the expression of selected genes across fibrosis stages (0–4) in two liver RNA-seq validation datasets. Spearman correlation coefficients (rho) indicate the strength of association between gene expression and fibrosis progression, while p-values from Kruskal-Wallis tests denote statistical significance.

## Supplemetary Methods

### Bulk transcriptomics data processing

#### Microarray data processing

An integrative analysis per chronic disease/fibrosis-affected organ system (kidney, heart, and liver) was conducted following the previously established pipeline (51). In brief, raw files of each GEO dataset were individually pre-processed applying background correction, quantile normalization and log transformation of expression values. Based on the above mentioned selection criteria, the microarray discovery cohort included: n=7 datasets for heart tissue (GSE57338, GSE52601, GSE42955, GSE26887, GSE79962, GSE81338 and GSE36961) n=4 datasets for kidney tissue covering n=2 datasets for glomeruli transcriptome (GSE104948 and GSE104066) and n=2 datasets for tubular transcriptome (GSE104954 and GSE200818), and samples from n=5 datasets for liver tissue (GSE152738, GSE77627, GSE103580, GSE89377 and GSE164760). Additionally, samples from n=4 microarray datasets for liver tissue, were selected as the initial validation cohort.

Individual datasets were merged per tissue, except for kidney tissue, where tubular and glomerular compartments were analyzed separately. This integration was achieved through common genes, and batch effect removal was performed using the ComBat algorithm (52). The batch effect correction was assessed using relative log expression and principal component analysis plots. All analyses were conducted using R software (R version 4.1.0, R Foundation for Statistical Computing, Vienna, Austria).

#### RNA-seq data processing

Two GEO single-end RNA-seq datasets of liver tissue (GSE193084 and GSE162694) from the Illumina platform were considered as a second validation cohort and analyzed individually. Raw FASTQ files from each dataset were downloaded from the GEO repository and aligned to the human reference genome sequence hg38 using STAR with default parameters. Summary counts of the number of aligned reads overlapping each annotated gene were obtained using the R package Rsubread with the featureCounts function.

### Single-cell data retrival

Single-cell RNA sequencing datasets were retrieved through a PubMed search using the keywords “liver fibrosis,” “NAFLD,” “NASH,” “cirrhosis,” and “hepatitis,”. Studies with available metadata detailing fibrosis stage and with public access to raw or processed snRNA-seq data were included. Those datasets involving infectious diseases or liver cancer were not considered for the analysis. To avoid integration bias and ensure methodological consistency across studies, most of which employed snRNA-seq, only single-nucleus RNA sequencing datasets were included, while single-cell RNA sequencing datasets were excluded.

### Single-nuclei transcriptomics data processing and batch effect correction

Data processing and downstream analysis were performed using the Python package Scanpy (v1.11.0). Raw count matrices were obtained and loaded into Scanpy objects for quality control filtering. Cells expressing fewer than 500 or more than 5,000 genes, or exhibiting over 10% mitochondrial transcript content, were excluded. After quality filtering, the resulting AnnData objects from individual samples were merged into a single dataset for subsequent integration and analysis. A total of 249,088 high-quality cells with 17,684 genes were retained for subsequent analysis. To perform dimensionality reduction, batch correction, and clustering, the top 4,000 highly variable genes across batches were identified.

The batch effect arising from diverse platforms and protocols was subsequently removed with the with the BBKNN algorithm using the Python package BBKNN (v1.6.0) using 50 PCs. Clustering in the integrated object was performed using the Leiden algorithm with parameter resolution = 3, and clusters were visualized using Uniform Manifold Approximation and Projection (UMAP) with parameters min_dist = 0.1, and spread = 1.4.

