## Supplementary material for "Pan-fibrotic gene expression signature in major chronic diseases by integrative bulk and single-cell transcriptomic analyses": Tables S1 to S8;Figs. S1 to S7

Mayra Alejandra Jaimes Campos *et al.*

***Corresponding author. Dr. Agnieszka Latosinska**

**This PDF file includes:**

Tables S1 to S8
Figs. S1 to S7

*Supplementary Tables*

| Dataset accession | Assay technology | Original number of samples | Excluded | Exclusion criteria | Sample type | Final number of samples | Cases | Disease/condition | Controls | Control samples | Reference (Pubmed ID) |
|---|---|---|---|---|---|---|---|---|---|---|---|
| **GSE57338** | Affymetrix Human Gene 1.1 ST Array | 313 | 4 | Age below 18yrs | Left ventricular tissue | 309 | 176 | Idiopathic dilated cardiomiopathy, ischemic cardiomiopathy | 133 | Unused heart donors | PMID: 25528681 |
| **GSE52601** | Illumina HumanHT-12 V4.0 | 24 | 5 | Age below 18yrs | Left ventricular tissue | 19 | 16 | Ischemic cardiomiopathy, dilated cardiomiopathy | 3 | Non-failing heart donors | PMID: 25012294 |
| **GSE42955** | Affymetrix Human Gene 1.0 ST Array | 29 | 0 | - | Left ventricular tissue | 29 | 24 | Ischemic cardiomiopathy, dilated cardiomiopathy | 5 | Unused heart donors | PMID: 24339868 |
| **GSE26887** | Affymetrix Human Gene 1.0 ST Array | 24 | 0 | - | Left ventricular tissue | 24 | 19 | Diabetic and non-diabetic heart failure | 5 | Non-failing heart donors | PMID: 22427379 |
| **GSE79962** | Affymetrix Human Gene 1.0 ST Array | 51 | 11 | Sepsis | Left ventricular tissue | 31 | 20 | Ischemic heart disease, dilated cardiomyopathy | 11 | Non-failing heart donors | PMID: 28067713 |
| **GSE81338** | Illumina HumanHT-12 V4.0 | 30 | 16 | Samples from right ventricle and replicates from the same patient in left ventricle | Left ventricular tissue | 14 | 14 | Dilated cardiomyopathy | - | - | PMID: 27417303, PMID: 27699191 |
| **GSE36961** | Illumina HumanHT-12 V3.0 | 145 | 16 | Age below 18yrs | Left ventricular tissue | 129 | 93 | Hypertrophic Cardiomyopathy | 36 | Unused heart donors | NA |

***Table S1. Selected heart tissue microarray datasets.***

| Dataset accession | Assay technology | Original number of samples | Excluded | Exclusion criteria | Sample type | Final number of samples | Cases | Disease/ condition | Controls | Control samples | Reference (Pubmed ID) |
|---|---|---|---|---|---|---|---|---|---|---|---|
| **GSE104954** | Affymetrix Human Genome U133 Plus 2.0 | 195 | 112 | Tumor nephrectomy, thin membrane disease, systemic lupus erythematosus, focal segmental glomerulosclerosis, minimal change disease, ANCA-associated vasculitis, | Microdissection on kidney biopsies (tubular compartment) | 83 | 62 | Chronic kidney disease (Diabetic nephropathy, hypertensive nephropathy, IgA nephropathy). | 21 | Living donor biopsies | PMID: 29724730 |
| **GSE200818** | Affymetrix Human Gene 2.1 ST | 193 | 132 | Focal and Segmental Glomerulosclerosis, Minimal Change Disease | Microdissection on kidney biopsies (tubular compartment) | 61 | 56 | Chronic kidney disease (Glomerular Disease (IgA nephropathy)). | 5 | Living donor biopsies | PMID: 23393107 and PMID: 23325076; PMID: 28339906 |
| **GSE104948** | Affymetrix Human Genome U133 Plus 2.0 | 196 | 121 | Tumor nephrectomy, thin membrane disease, systemic lupus erythematosus | Microdissection on kidney biopsies (glumeruli compartment) | 75 | 54 | Chronic kidney disease (Diabetic nephropathy, hypertensive nephropathy, IgA nephropathy). | 21 | Living donor biopsies | PMID: 29724730 |
| **GSE104066** | Affymetrix Human Gene 2.1 ST | 76 | 48 | Membranous Nephropathy, Focal and Segmental Glomerulosclerosis, Minimal Change Disease | Microdissection on kidney biopsies (glumeruli compartment) | 28 | 22 | Chronic kidney disease (Glomerular Disease (IgA nephropathy)). | 6 | Living donor biopsies | PMID: 30301568 |

***Table S2. Selected kidney tissue microarray datasets from tubuli and glomeruli.***

| Dataset accession | Assay technology | Original number of samples | Excluded | Exclusion criteria | Final number of samples | Sample type | Cases | Disease/ condition | Control¥ | Control samples | Reference (Pubmed ID) |
|---|---|---|---|---|---|---|---|---|---|---|---|
| **GSE152738** | Affymetrix Human Genome U133 Plus 2.0 | 58 | - | - | 58 | Liver specimen | - | - | 58 | Healthy liver donors | PMID: 33888584 |
| **GSE77627** | Illumina HumanHT-12 WG-DASL V4.0 R2 | 54 | 18 | Idiopathic noncirrhotic portal hypertension | 36 | Liver specimen | 22 | Cirrhosis | 14 | Histologically normal livers | PMID: 34052252 |
| **GSE103580** | Affymetrix Human Genome U219 | 86 | 19 | NAFLD and NASH without information on fibrosis stage | 67 | Tissue biopsies | 67 | Cirrhosis | 6 | Histologically normal livers | PMID: 29158192 |
| **GSE89377** | Illumina HumanHT-12 V3.0 | 107 | 62 (+20*) | HCC, dysplastic nodules (*including those used for validation) | 25 | Liver specimen | 12 | Cirrhosis | 13 | Healthy liver donors | PMID: 29059470; PMID: 35205612 |
| **GSE164760** | Affymetrix Human Genome U219 | 170 | 127 | Adjacent non-tumor NASH, HCC and NASH without information on fibrosis stage | 14 | Liver specimen | 8 | Cirrhosis | 6 | Healthy liver donors | PMID: 33992698 |

¥The same set of controls (GSE152738, GSE77627, GSE103580, GSE89377, GSE164760, GSE84044) were used for the microarray discovery and validation cohorts.

***Table S3. Selected liver tissue microarray datasets.***

| Dataset accession | Assay technology | Original number of samples | Excluded | Exclusion criteria | Sample type | Final number of samples | Cases | Disease/ Condition | Control[¥] | Control samples | Reference (Pubmed ID) |
|---|---|---|---|---|---|---|---|---|---|---|---|
| GSE89377 | Illumina HumanHT-12 V3.0 | 107 | 62 (+12*) | HCC and dysplastic nodules (*including those used for discovery) | Liver specimen | 33 | 20 | Chronic hepatitis | 13 | Healthy liver donors | PMID: 29059470; PMID: 35205612 |
| GSE73634 | Affymetrix Human Gene 2.0 ST | 48 | 12 | Post-treatment with simtuzumab | Tissue biopsies | 12 | 12 | NASH | 0 | - | PMID: 27232579 |
| GSE84044 | Affymetrix Human Genome U133 Plus 2.0 | 124 | | | Tissue biopsies | 124 | 81 | Chronic hepatitis | 43 | NAFLD fibrosis stage 0 | PMID: 28262670 |
| GSE49541 | Affymetrix Human Genome U133 Plus 2.0 | 72 | | | Tissue biopsies | 72 | 72 | NAFLD | 0 | - | PMID: 23913408 and PMID: 23916847 |

[¥]The same set of controls (GSE152738, GSE77627, GSE103580, GSE89377, GSE164760, GSE84044) were used for the microarray discovery and initial validation cohorts.

***Table S4. Selected microarray initial validation cohort datasets.***

| Dataset accession | Assay technology | Original number of samples | Excluded | Exclusion criteria | Sample type | Final number of samples | Cases | Disease/condition | | | | Controls | Control samples | Reference (Pubmed ID) |
|---|---|---|---|---|---|---|---|---|---|---|---|---|---|---|
| | | | | | | | | NASH | | | | | | |
| | | | | | | | | **Stage 1** | **Stage 2** | **Stage 3** | **Stage 4** | | | |
| **GSE193084** | Illumina NextSeq 500 | 271 | 0 | - | Tissue biopsies | 271 | 259 | 66 | 84 | 71 | 38 | 12 | Histologically normal livers and NAFLD fibrosis stage 0 | PMID: 29724730 |
| | | | | | | | | NASH | | | | | | |
| | | | | | | | | **Stage 1** | **Stage 2** | **Stage 3** | **Stage 4** | | | |
| **GSE162694** | Illumina HiSeq 3000 | 143 | 0 | - | Tissue biopsies | 143 | 77 | 30 | 27 | 8 | 12 | 66 | NAFLD fibrosis stage 0 | PMID: 34508113 |

***Table S5. Independent RNA-seq second validation cohort.***

| Dataset accession | Assay technology | Original number of samples | Excluded | Exclusion criteria | Sample type | Final number of samples | Cases | Disease/condition | Stage 1 | Stage 2 | Stage 3 | Stage 4 | Controls | Control samples | Reference (Pubmed ID) |
|---|---|---|---|---|---|---|---|---|---|---|---|---|---|---|---|
| **GSE185477** | Illumina HiSeq 2500 | 4 | - | - | Liver specimen | 4 | 0 | - | - | - | - | - | 4 | Neurologically deceased donor liver acceptable for liver transplantation. | PMID: 34792289 |
| **GSE244832** | Illumina NextSeq 500 | 18 | - | - | Liver specimen | 18 | 9 | NAFLD, NASH, cirrhosis | - | 1 | 4 | 4 | 9 | Histologically normal livers and NAFLD fibrosis stage 0 | PMID: 39522884 |
| **GSE210077** | Illumina NextSeq 500 | 35 | 29 | No description of fibrosis stage | Liver specimen | 6 | 3 | No specified | - | 1 | 1 | 1 | 3 | Healthy liver donors | PMID: 39747812 |
| **GSE202379** | Illumina NovaSeq 6000 | 47 | 5 | End stage | Liver specimen and tissue biopsies | 42 | 37 | | 9 | 12 | 12 | 4 | 5 | Histologically normal livers and NAFLD fibrosis stage 0 | PMID: 38778114 |

*Table S6. Independent snRNA-seq validation cohort.*

| Ishak score | Scheuer system and METAVIR system | Pathology Committee of the NASH Clinical Research Network | Not specified | Unified system |
|---|---|---|---|---|
| 0 (no fibrosis) | F0 (no fibrosis-“mild“) | Stage 0 (no fibrosis) | | No fibrosis |
| 1 (mild fibrosis) | F1 (mild to moderate fibrosis) | Stage 1 (mild fibrosis) | low-grade fibrosis of chronic hepatitis | Low fibrosis |
| 2(mild to moderate fibrosis) | Either or between F1 and F2 | Either or between stage 1 and 2 (mild to moderate fibrosis) | | Moderate fibrosis |
| 3 (moderate fibrosis) | F2 (moderate fibrosis9 | Stage 2 (moderate fibrosis) | | Moderate fibrosis |
| 4 (moderate to severe fibrosis) | F3 (severe fibrosis) | Stage 3 (severe fibrosis) | high-grade fibrosis of chronic hepatitis | Severe fibrosis |
| 5 (incomplete cirrhosis) | Either or between F3 or F4 (severe fibrosis or cirrhosis) | Either or between stage 3 or 4 (severe fibrosis or cirrhosis) | | Severe fibrosis |
| 6 (cirrhosis) | F4 (cirrhosis) | Stage 4 (cirrhosis) | | Severe fibrosis |

*Table S7. Unification of the different systems describing the degree of liver fibrosis in the microarray validation cohort.*

| Cell type | Canonical markers |
|---|---|
| Hepatocytes | AL391117.1, ADRA1A, ACSM2B, HNF4A, CYP2A6, ABCC2, ASGR1 |
| Monocytes | FCGR3A, MS4A7,CD163, CSF1R |
| T cells | IL7R, CD96, CD247 |
| B cells | MS4A1 |
| Cholangiocytes | CTNND2, DCDC2, CFTR, SLC12A2 |
| Endothelial | ST6GALNAC3, PTPRB, PECAM1, FLT1, EGFL7, ERG |
| Stellate cells | PRKG1, LAMA2, ARHGAP10, ADAMTS9-AS2, PDGFRA, ADAMTSL2 |

***Table S8. Canonical markers for cell cluster annotation.***

### Supplementary Figures

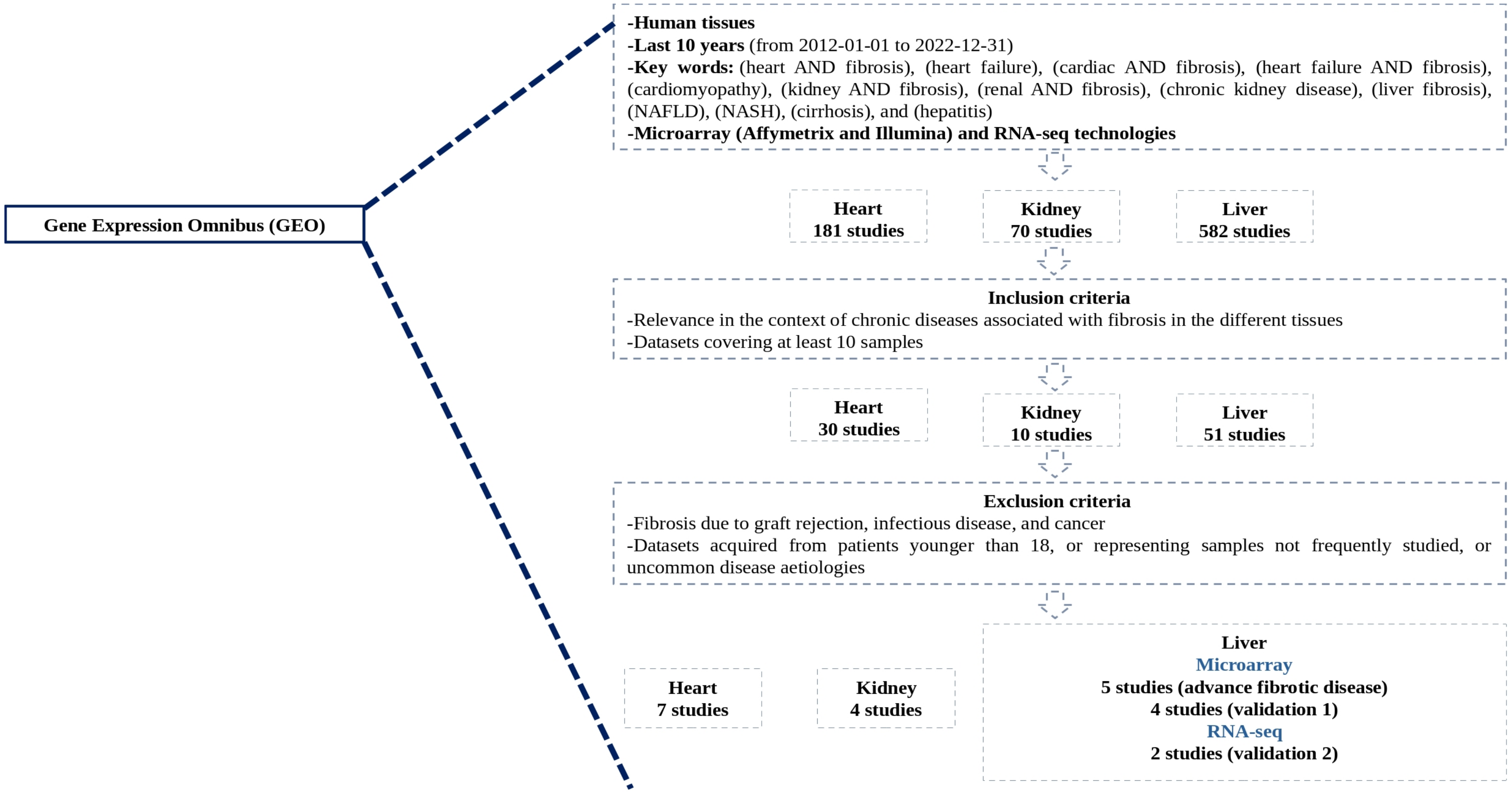


*Figure S1.* ***Overall workflow for the retrieval and curation of microarray, and RNAseq publicly available data representing fibrotic and non-fibrotic conditions.***

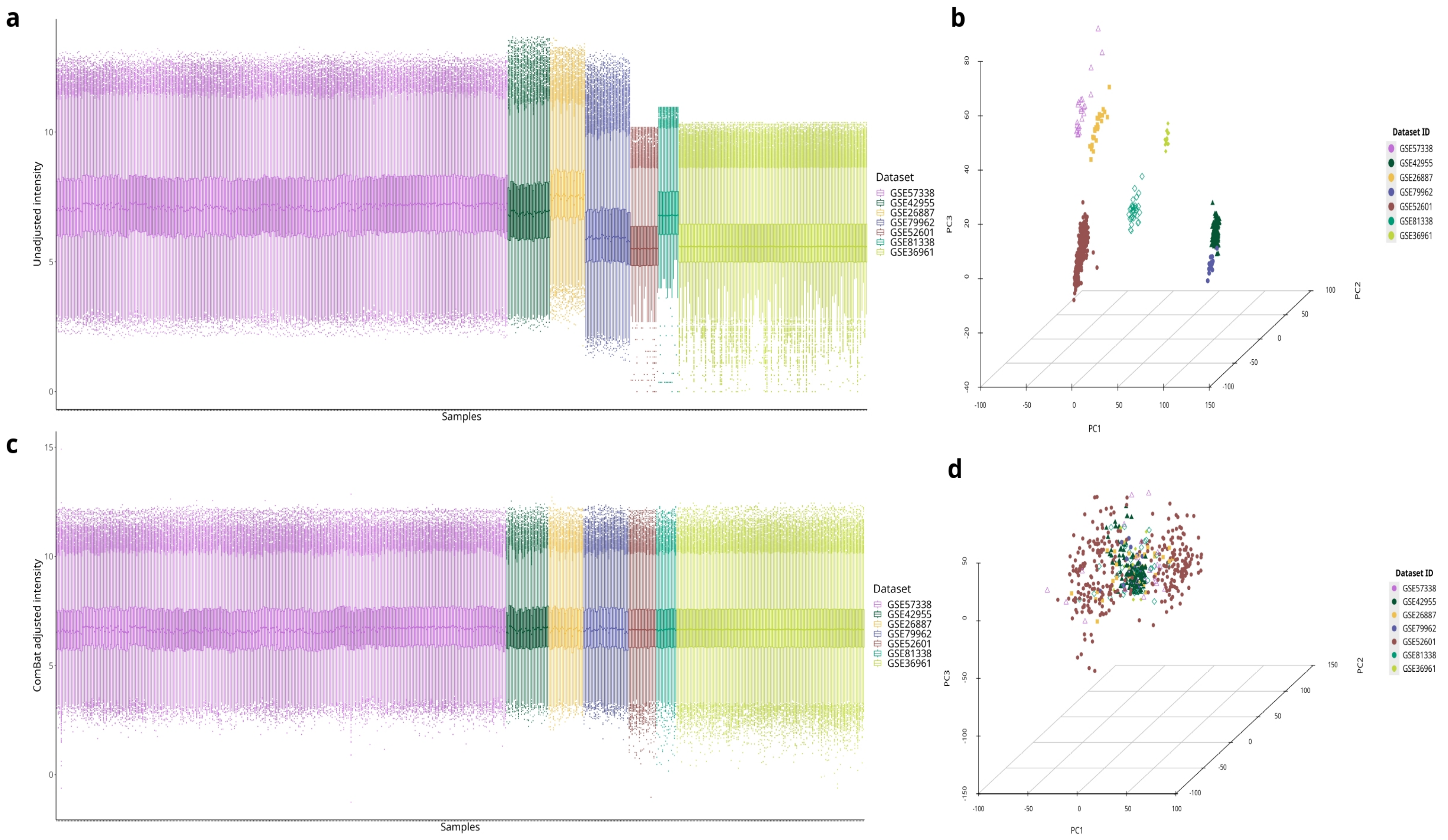


***Figure S2. Batch effect correction of heart microarray datasets (discovery).*** *a-b. Box plots and PCA plots of unadjusted expression data, showing dataset-specific variability. c-d. Box plots and PCA plots after ComBat adjustment, illustrating successful batch effect correction with improved sample distribution and reduced dataset-specific clustering.*

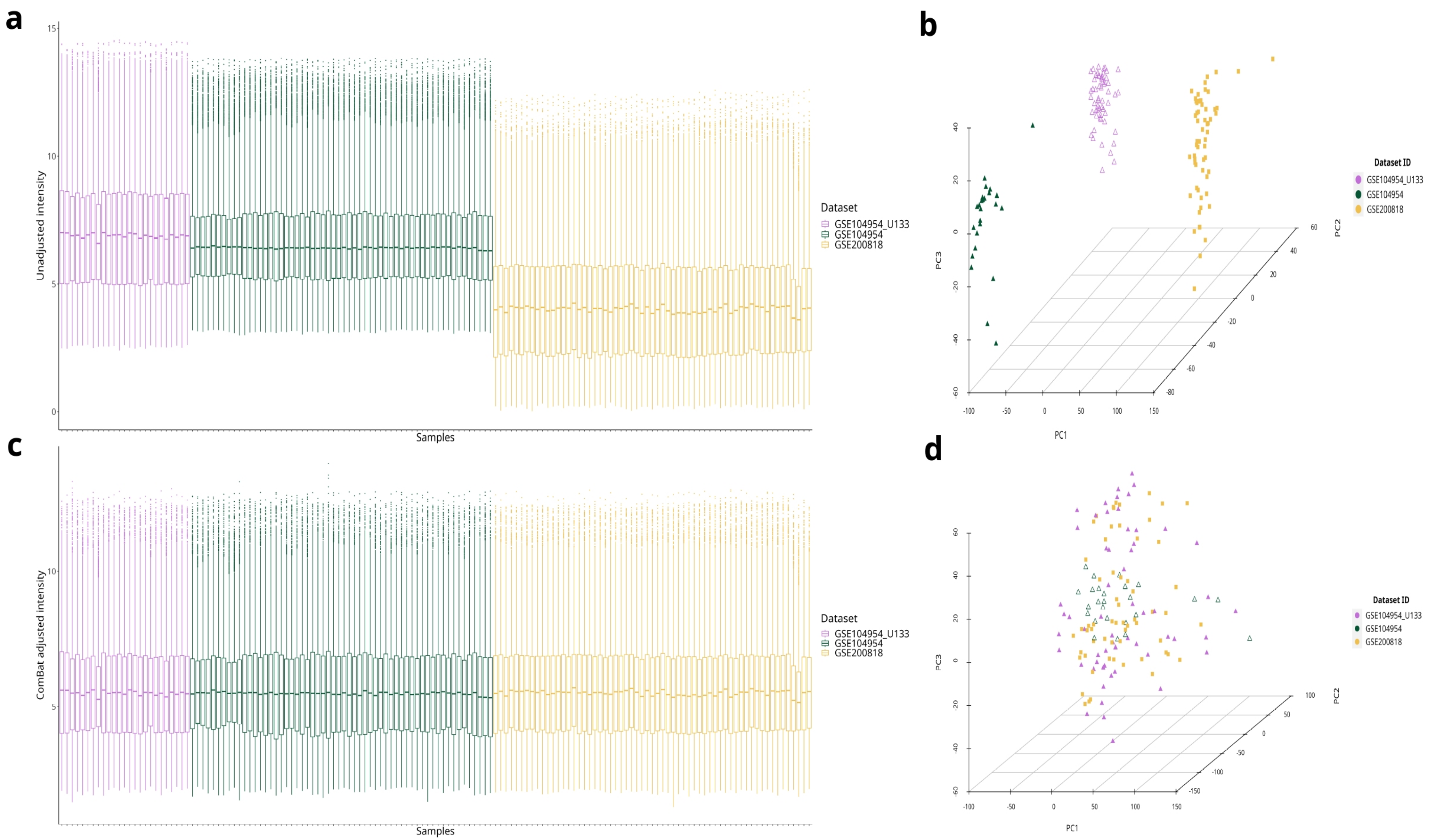


***Figure S3. Batch effect correction of kidney tubuli microarray datasets (discovery).*** *a-b. Box plots and PCA plots of unadjusted expression data, showing dataset-specific variability. c-d. Box plots and PCA plots after ComBat adjustment, illustrating successful batch effect correction with improved sample distribution and reduced dataset-specific clustering.*

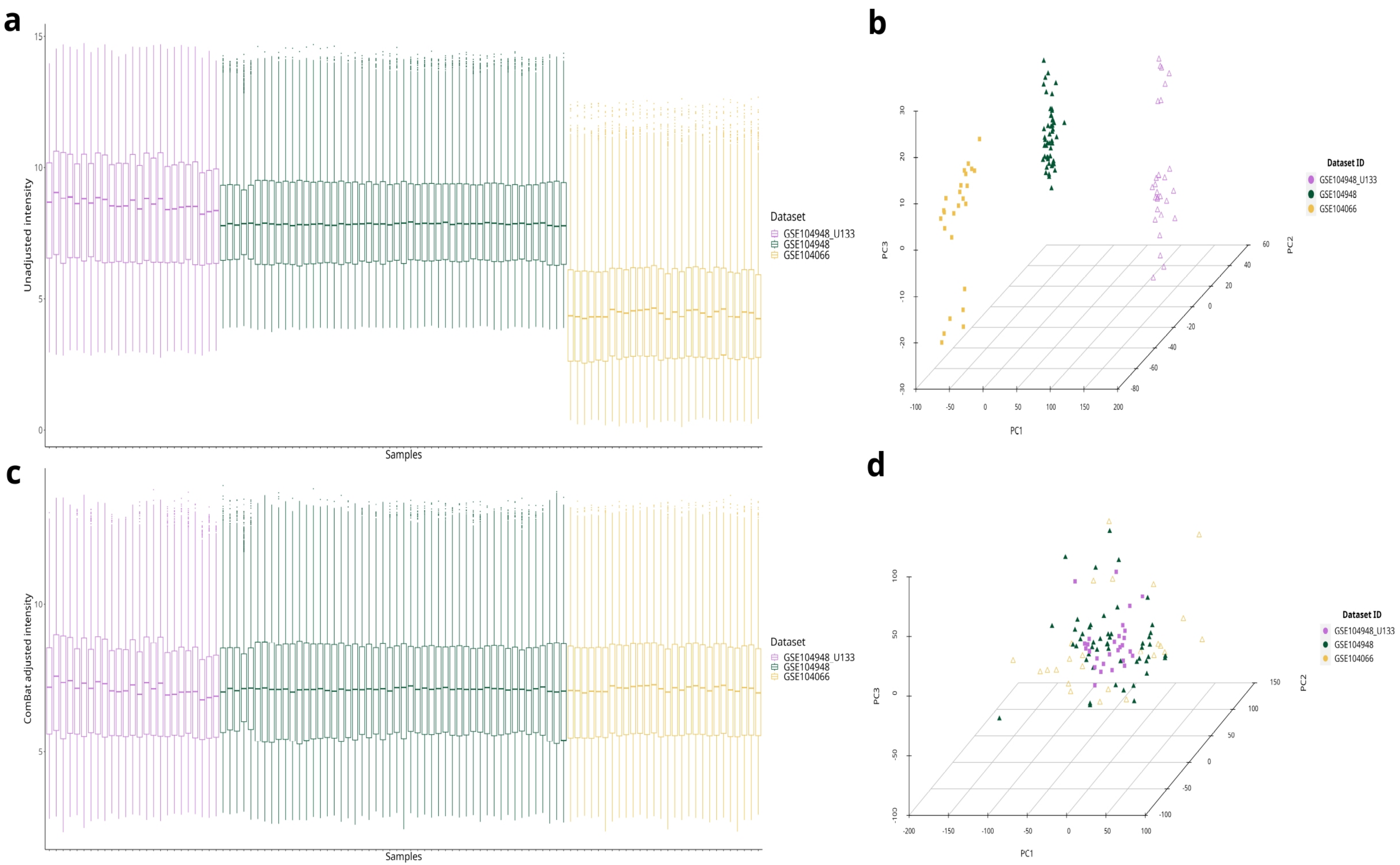


***Figure S4. Batch effect correction of kidney glomeruli microarray datasets (discovery).*** *a-b. Box plots and PCA plots of unadjusted expression data, showing dataset-specific variability. c-d. Box plots and PCA plots after ComBat adjustment, illustrating successful batch effect correction with improved sample distribution and reduced dataset-specific clustering.*

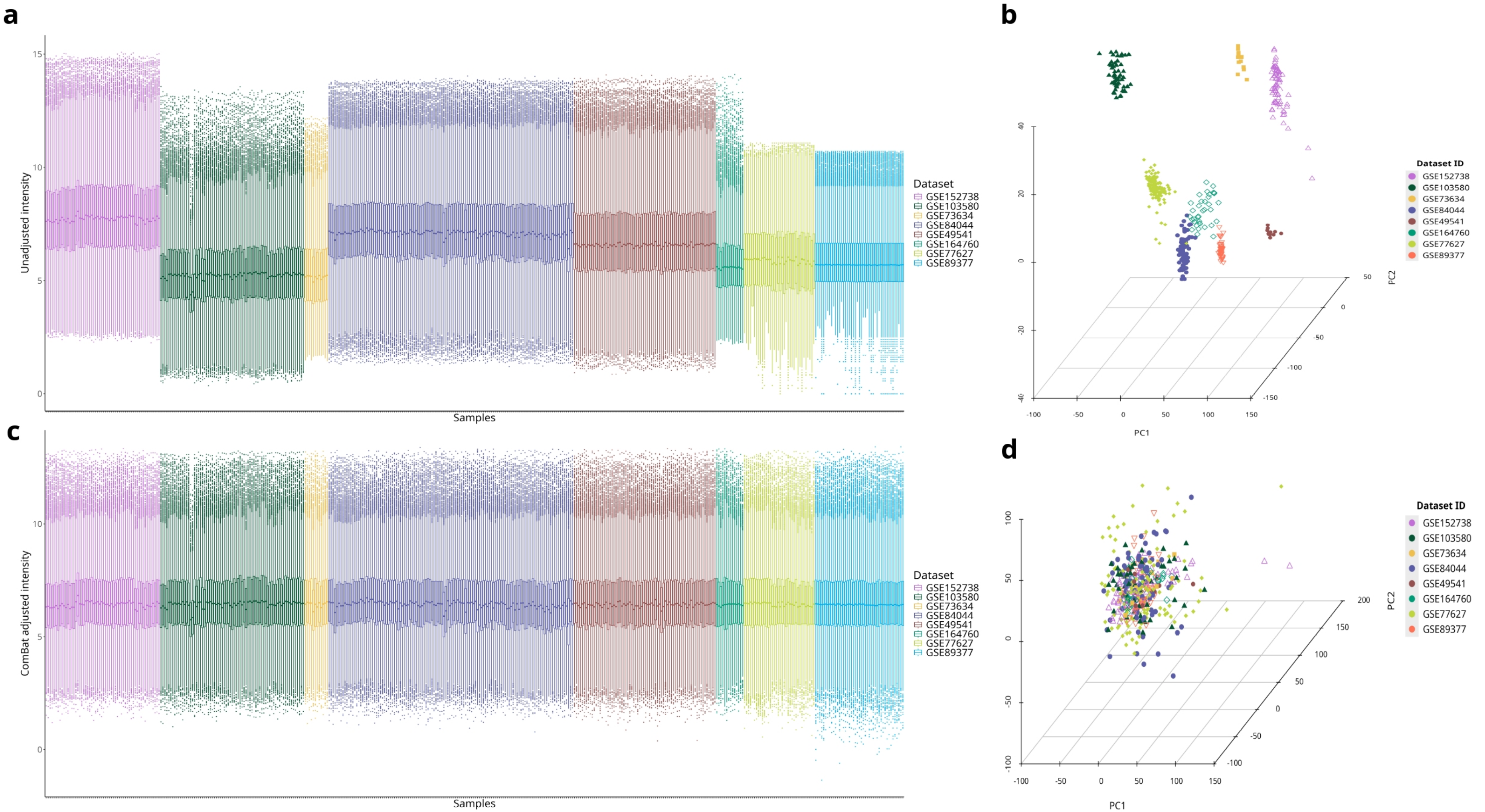


***Figure S5. Batch effect correction of liver microarray datasets (discovery + initial validation cohorts).*** *a-b. Box plots and PCA plots of unadjusted expression data, showing dataset-specific variability. c-d. Box plots and PCA plots after ComBat adjustment, illustrating successful batch effect correction with improved sample distribution and reduced dataset-specific clustering.*

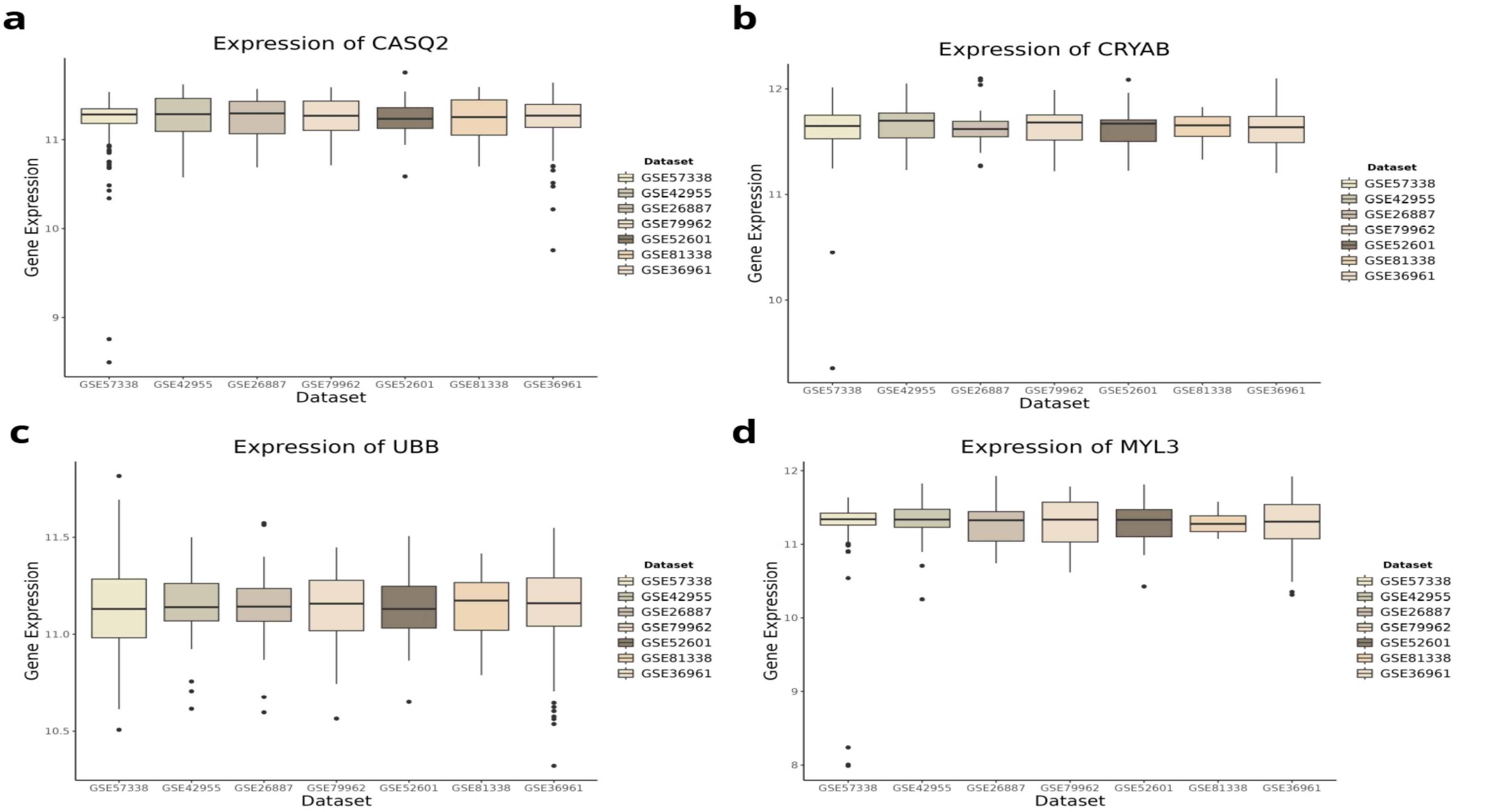


***Figure S6. ComBat-adjusted expression of housekeeping genes across individual heart microarray datasets.*** *a-d. Box plots showing the high and consistent mean expression levels of selected housekeeping genes after batch effect correction using the ComBat algorithm.*

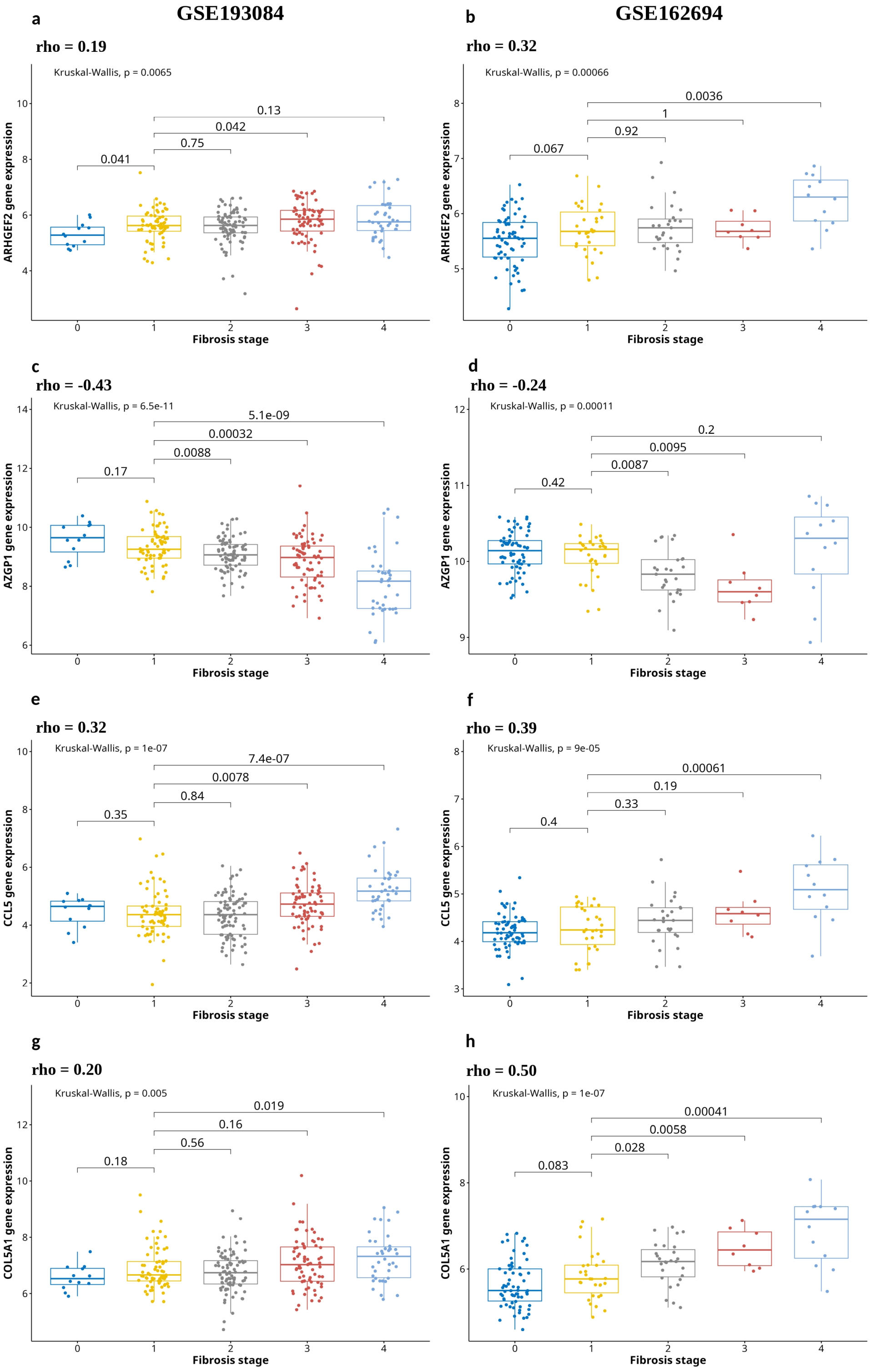


***Figure S7. Box plots illustrating the association of hallmark elements of the pan-fibrotic gene-expression signature, with fibrosis progression in the RNA-seq liver validation datasets.*** *a-h. Box plots showing the expression of selected genes across fibrosis stages (0–4) in two liver RNA-seq validation datasets. Spearman correlation coefficients (rho) indicate the strength of association between gene expression and fibrosis progression, while p-values from Kruskal-Wallis tests denote statistical significance.*
